# Long Sleep and Insomnia are Associated with Reduced Global Cognitive Function across Diverse Populations

**DOI:** 10.64898/2026.10.19.26365906

**Authors:** Annah B. Wyss, Yu Zhang, Michael R. Brown, Jan Bressler, Charles DeCarli, Myriam Fornage, Hector M. González, Xiuqing Guo, Susan R. Heckbert, Timothy M. Hughes, Carmen R. Isasi, Sonya S. Kaur, Melissa Lamar, Peter Y. Liu, Pamela L. Lutsey, Alanna C. Morrison, Qibin Qi, Alberto R. Ramos, Susan Redline, Jerome I. Rotter, Chloé Sarnowski, Daniela Sotres-Alvarez, Wassim Tarraf, Kent D. Taylor, Meredith J. Wallace, Alexis C. Wood, Tamar Sofer

## Abstract

**Background:** Although sleep disorders are generally associated with cognitive decline, definitions of sleep conditions and cognitive function vary greatly across studies. Therefore, we evaluated associations between consensus definitions of sleep and cognitive outcomes across multiple studies.

**Methods:** Consensus sleep measures were ascertained at baseline visits in three cohort studies (HCHS/SOL, MESA and ARIC, N=11,563): short and long sleep (≤6hours and ≥9hours), insomnia (Women’s Health Initiative Insomnia Rating Scale≥10 or ≥6 for 3 questions), daytime sleepiness (Epworth Sleepiness Scale≥11) and obstructive sleep apnea (OSA, Apnea-Hypopnea Index≥ 15). Cognitive measures were ascertained at baseline and follow-up visits in each study and used to calculate harmonized outcomes: global cognitive function (GCF) at baseline, change in GCF between baseline and follow-up, poor GCF at baseline (≥1SD below the median), and incident poor GCF at follow-up. Study-specific analyses were adjusted for age, sex, center, education, race/ethnicity, *APOE4* and language of cognitive assessments as needed. Results were meta-analyzed across studies.

**Results:** Long sleep was consistently associated with lower baseline GCF (β=-0.19, SE=0.03, p=3.78x10^-14^, FDR p=7.56x10^-13^, p-heterogeneity=0.78) and increased odds of poor baseline GCF (OR=1.77, 95% CI=1.49-2.09, p=2.96x10^-11^, FDR p=2.96x10^-10^, p-heterogeneity=0.22) across cohorts. Insomnia was also suggestively associated with lower baseline GCF (β=-0.051, SE=0.02, p=0.01, FDR p=0.07, p-heterogeneity=0.005) and increased odds of poor baseline GCF (OR=1.22, 95% CI=1.06-1.41, p=6.76x10^-3^, FDR p=0.05, p-heterogeneity=0.03). Sleep measures were not associated with change in cognitive function.

**Discussion:** Associations of long sleep and insomnia with harmonized cross-sectional GCF across three large, multiethnic cohorts suggest that cognitive function may be associated with distinct sleep phenotypes.

## Introduction

Age-related cognitive decline is an important public health concern ^1^. Several factors can influence cognitive health, including genetic, demographic and lifestyle measures ^1^. Studying the relationship between modifiable risk factors and cognitive measures is important to develop early interventions to slow cognitive decline or mitigate the onset of conditions such as Alzheimer’s disease and related dementias. Given the important role of sleep in cognitive aging among older adults ^2^, improved characterization of this relationship could facilitate identification and prioritization of phenotypes for public health intervention.

Previous studies on the associations between insomnia, excessive daytime sleepiness (EDS) and obstructive sleep apnea (OSA) with cognitive performance have reported mixed results ^2–5^. Likewise, several studies of sleep duration reported U-shaped risk curves with both short and long sleep durations associated with reduced cognitive function, while other studies reported associations for only long sleep or only short sleep ^2,6–8^. Variation in results likely reflects differences in how sleep and cognitive measures were ascertained or defined. While polysomnography (PSG) is the gold standard for measuring many attributes of sleep, it is resource-intensive and can be influenced by night-to-night and scoring variability ^9,10^. Self-reported sleep measures obtained by questionnaire are more commonly available, but questions often vary across studies ^11,12^. Cognitive function can be measured using a variety of assessments, including broad screening instruments such as Mini-Mental Status Exam (MMSE) or Montreal Cognitive Assessment (MoCA) as well as specific tests or larger batteries covering neuropsychological domains such as memory, language, executive function, abstract reasoning, attention, and visuospatial skills ^13^. Even among studies that use similar cognitive assessment tools, examinations can vary in terms of specific questions or time allowed to complete tasks ^14^. For this reason, some studies standardize and combine data from cognitive assessments to produce composite global cognitive function scores ^15–17^. Previous studies of the association between sleep and cognition have utilized a vast array of combinations of sleep and cognitive measures ^3–6^. Furthermore, some previous studies examined the relationship between sleep and cognition cross-sectionally considering only prevalent measures, while other studies prospectively considered associations with incident cognitive measures ^3–6^.

Given differences in results and methods across previous studies, there is a need to better harmonize data across studies and understand which sleep-cognitive function associations are consistent across populations. While some sleep phenotypes may have population-specific associations with cognitive function (e.g., due to systematic differences in sociodemographic measures and environmental exposures between populations), identification of sleep phenotypes that are consistently associated with cognitive outcomes suggest associations that are robust to confounders, measurement variability, and population-specific susceptibilities. Therefore, the present study seeks to extend previous research by utilizing and harmonizing data from 12,811 individuals from three longitudinal cohort studies with available sleep and cognitive measures: The Hispanic Community Health Study/Study of Latinos (HCHS/SOL), The Multi-Ethnic Study of Atherosclerosis (MESA) and The Atherosclerosis Risk in Communities (ARIC) Study. Each of these cohorts represents a unique sample of the U.S. population. We harmonized cognitive measures to generate comparable global cognitive function variables, and study associations with well-defined and generalizable sleep measures across cohorts.

## Methods

### Cohort Studies

#### The Hispanic Community Health Study/Study of Latinos

The Hispanic Community Health Study/Study of Latinos (HCHS/SOL) is a longitudinal study of 16,415 individuals of Hispanic/Latino origin (Cuban, Puerto Rican, Dominican, Mexican, Central and South American) ^18,19^. Individuals were aged 18 to 74 years at enrollment, with oversampling of individuals aged 45 to 74 years. A two-stage probability sampling design based on census-blocks groups and household addresses was employed to enroll individuals from four geographic sites in the US (Miami, San Diego, Chicago, and the Bronx area of New York). To account for the differences in probability sampling, analyses were adjusted for sampling weights and accounted for clustering and stratification.

Self-reported sleep measures were ascertained at the HCHS/SOL baseline visit (2008-2011) and included sleep duration, insomnia and excessive daytime sleepiness (Table 1). Sleep duration was computed as a weighted average of weekend and weekday sleep based on the questions “what time do you usually go to bed?” and “what time do you usually wake up?” on weekdays and weekends. Insomnia was based on the Women’s Health Initiative Insomnia Rating Scale (WHIIRS) ^20^ and excessive daytime sleepiness was based on the Epworth Sleepiness Scale (ESS) ^21^. Obstructive sleep apnea was also ascertained at baseline based on self-administered, in-home sleep tests using an ARES Unicorder 5.2 (B-Alert, Carlsbad, CA) from which the respiratory event index (REI) was calculated as the number of respiratory events (defined as ≥50% reduction in airflow and ≥3% desaturation for ≥10 seconds) per sleep hour ^22^.

**Table 1.**
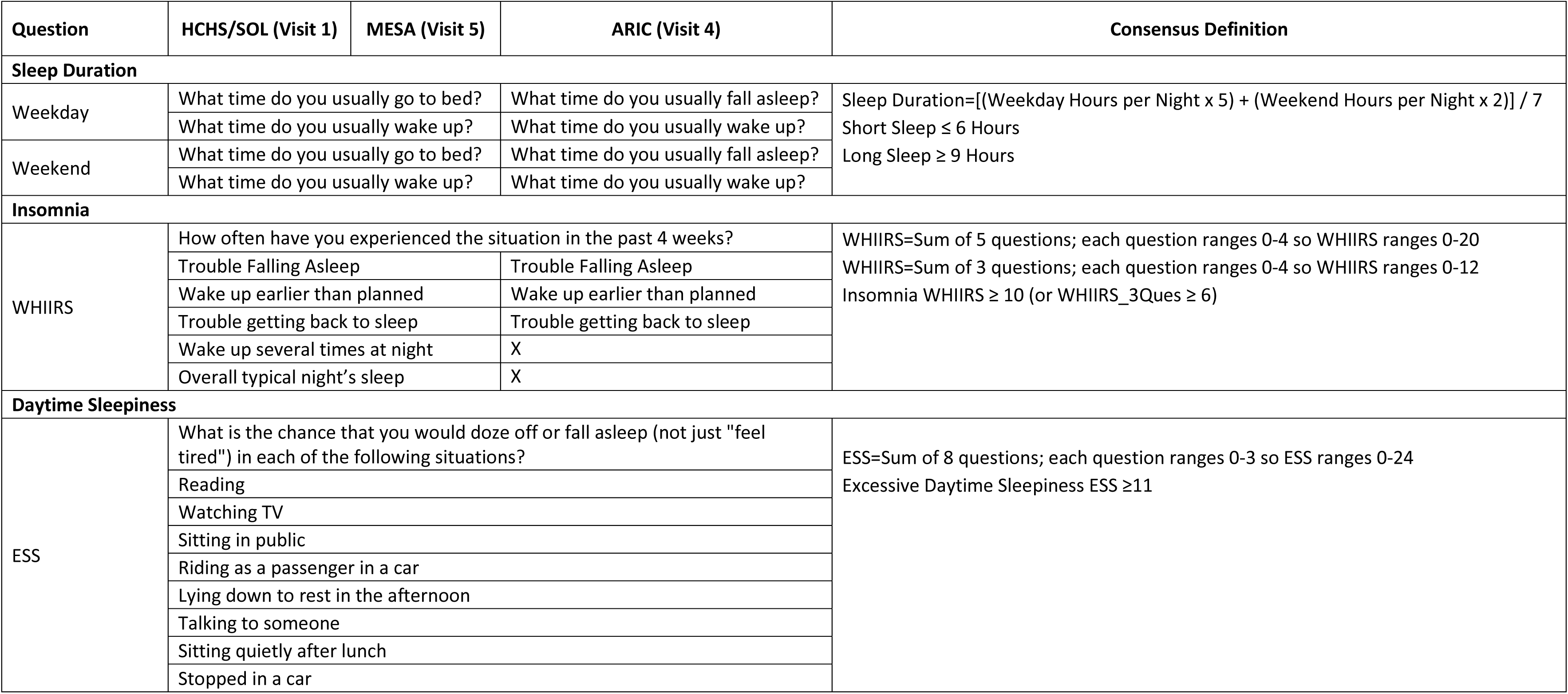
Sleep Questions from HCHS/SOL (Visit 1), MESA (Visit 5), and ARIC (Visit 4) and Harmonized Definitions.

Cognitive measures were ascertained at HCHS/SOL baseline visit (2008-2011) and in an ancillary study, the Study of Latinos-Investigation of Neurocognitive Aging (SOL-INCA; N=6,377; 2015-2018), that occurred around the same time as the HCHS/SOL second follow-up visit ^23^. Cognitive exams included a six-item screener (SIS), the digit symbol substitution test (DSST), word fluency test (WFT), and a brief Spanish English verbal learning test (B-SEVLT) (Table 2). While additional cognitive tests were available in the follow-up visit, we only used those available in both visits. Participants could complete cognitive exams in their language of preference, either English (80%) or Spanish (20%). 8,458 individuals with sleep, cognitive and complete covariate (base model) data were included in the present analysis.

**Table 2.** Cognitive Function Tests Available in HCHS/SOL (Visit 1 and 2), MESA (Visit 5 and 6), and ARIC (Visit 4 and 5) and Harmonized Global Cognitive Function Definitions.

| Cognitive Function Test | Cognitive Domain | HCHS/SOL (Visit 1 and 2) | MESA (Visit 5 and 6) | ARIC (Visit 4 and 5) | Consensus Definition |
| --- | --- | --- | --- | --- | --- |
| Digit Symbol Substitution Test (DSST) | Executive Function, Speed of Processing | Yes | Yes | Yes | 1) Estimated sample means and standard deviations (SDs) for each test at baseline<br>2) Standardized test scores at baseline and follow-up by subtracting the test-specific baseline mean and dividing by the baseline SD<br>3) Baseline global cognitive function (GCF) computed from principal components analysis (PCA) using the baseline standardized test-specific scores available within each cohort. GCF defined as standardized first PC (PC1).<br>4) Follow-up GCF computed from the follow-up standardized test-specific scores available within each cohort and weighted using baseline PC1 loadings (i.e., projection of baseline PC1 on follow-up data). GCF defined as this variable, standardized by baseline PC1 mean and SD.<br>5) GCF change was defined as the GCF value at follow-up minus the GCF value at baseline.<br>6) Poor GCF was defined as GCF > 1 SD below the median at baseline in each cohort. Incident poor GCF was defined as not meeting this criterion at baseline but meeting this criterion at follow-up. |
| Cognitive Abilities Screening Instrument (CASI) | Global | No | Yes | No |  |
| Six-Item Screener (SIS)<sup>a</sup> | Global | Yes | No | No |  |
| Word Fluency Test (WFT) | Language | Yes | No | Yes |  |
| Brief Spanish English Verbal Learning Test (B-SEVLT)<sup>b</sup> | Verbal Learning and Memory | Yes | No | No |  |
| Delayed Word Recall Test (DWRT)<sup>b</sup> | Verbal Learning and Memory | No | No | Yes |  |
| Digit Span Test | Episodic memory | No | Yes | No |  |

a. When analyzing individual cognitive function tests, an SIS variable was created from subset of CASI questions in MESA and meta-analyzed with the SIS from HCHS/SOL.
b. When analyzing individual cognitive function tests, B-SEVLT (Delayed Recall) from HCHS/SOL and DWRT from ARIC meta-analyzed since test similar.

#### The Multi-Ethnic Study of Atherosclerosis (MESA)

The Multi-Ethnic Study of Atherosclerosis (MESA) is a longitudinal study of 6,814 individuals from diverse racial and ethnic backgrounds (approximately 38% of participants are White, 28% Black/African American, 22% Hispanic, and 12% Asian) ^24,25^. Participants were initially enrolled between 2000 and 2002 at six sites in the United States (Forsyth County NC, New York City NY, Baltimore City and County MD, St. Paul MN, Chicago IL, Los Angeles County CA). Individuals were aged 45-84 and had no known cardiovascular disease at time of enrollment. Sleep measures were ascertained during visit 5 (2010-2013) and included sleep duration, insomnia and excessive daytime sleepiness based on the same questions as HCHS/SOL (Table 1). OSA was measured using PSG with the apnea-hypopnea index (AHI) defined as the number of respiratory events (apneas and hypopneas with ≥ 4% oxygen desaturation) per sleep hour ^26^.

Cognitive measures were from visit 5 (2010-2012) and visit 6 (2016-2018), referred to as baseline and follow-up visits for the purposes of this analysis. Cognitive measures included the cognitive abilities screening instrument (CASI), the digit symbol coding test (same as DSST but participants given 120 seconds instead of 90 seconds to complete task), and the digit span test (Table 2). Participants could complete cognitive exams in English (80%), Spanish (11%), or Chinese (9%). 1,511 individuals with sleep, cognitive and complete covariate (base model) data were included in the present analysis.

#### The Atherosclerosis Risk in Communities (ARIC) Study

The Atherosclerosis Risk in Communities (ARIC) study is a longitudinal study of 15,792 individuals of mostly European and African ancestry ^27^. Participants were enrolled between 1987 and 1989 from four US communities (northwestern suburbs of Minneapolis, MN; Washington County, MD; Forsyth County, NC; and Jackson, MS) and were aged 45-64 years at time of enrollment. Sleep measures were ascertained for 1,920 ARIC participants from the Minnesota and Maryland field centers who participated in the Sleep Heart Health Study (SHHS) which occurred around the time of ARIC visit 4 (1996-1998) ^28^. Participants in SHHS were predominantly of European ancestry. The SHHS Sleep Habits Questionnaire was used to elicit information about habitual sleep duration, insomnia, and excessive daytime sleepiness (Table 1). Sleep duration was computed from the questions “what time do you usually fall asleep?” and “what time do you usually wake up?” on weekdays and weekends. Insomnia was based on three of the five questions from the WHIIRS: “How often have you experienced the situation in the past 4 weeks…Trouble Falling Asleep? Wake up earlier than planned? Trouble getting back to sleep?” Excessive daytime sleepiness was based on the ESS. OSA was measured using PSG with the apnea-hypopnea index (AHI) defined as the number of respiratory events (apneas and hypopneas with ≥ 4% oxygen desaturation) per sleep hour ^29^.

Cognitive measures were ascertained as part of ARIC visit 4 and visit 5 (2011-2013), referred to as baseline and follow-up visits for the purposes of this analysis. Cognitive exams included a delayed word recall test (DWRT), the DSST, and a WFT ^29^ (Table 2). Participants with prevalent stroke at the baseline or follow-up visits or prevalent dementia at the follow-up visit were excluded prior to the generation of global cognitive function variables. 1,594 individuals with sleep, cognitive data and complete covariate (base model) were included in the present analysis.

#### Ethics Statement

The HCHS/SOL was approved by the institutional review boards (IRBs) at each field center, where all participants gave written informed consent, and by the Non-Biomedical IRB at the University of North Carolina at Chapel Hill to the HCHS/SOL Data Coordinating Center. All IRBs approving the HCHS/SOL study are: Non-Biomedical IRB at the University of North Carolina at Chapel Hill, Chapel Hill, NC; Einstein IRB at the Albert Einstein College of Medicine of Yeshiva University, Bronx, NY; IRB at Office for the Protection of Research Subjects (OPRS), University of Illinois at Chicago, Chicago, IL; Human Subject Research Office, University of Miami, Miami, FL; and Institutional Review Board of San Diego State University, San Diego, CA.

All MESA participants provided written informed consent, and the study was approved by the Institutional Review Boards at The Lundquist Institute (formerly Los Angeles BioMedical Research Institute) at Harbor-UCLA Medical Center, University of Washington, Wake Forest School of Medicine, Northwestern University, University of Minnesota, Columbia University, Johns Hopkins University, and Brigham and Women’s Hospital.

For ARIC, written informed consent was provided by all study participants and the study design and methods were approved by IRBs at each of the collaborating medical institutions: University of Mississippi Medical Center Institutional Review Board (Jackson Field Center); Wake Forest University Health Sciences Institutional Review Board (Forsyth County Field Center); University of Minnesota Institutional Review Board (Minnesota Field Center); and Johns Hopkins Bloomberg School of Public Health Institutional Review Board (Washington County Field Center).

All methods and analyses were carried out in accordance with human subject research guidelines and regulations. This work was approved by the Beth Israel Deaconess Medical Center Committee on Clinical Investigations.

#### Consensus Sleep Measures

In a previous publication ^30^, we evaluated several consensus definitions of self-reported short and long sleep duration, insomnia and excessive daytime sleepiness across HCHS/SOL, MESA and ARIC by estimating associations with sleep trait-specific polygenic scores (PGS), with hypertension, and with the Digit Symbol Substitution Test (DSST). Based on statistical significance, consistency across studies, and sample sizes of these prior association analyses, we adopted the following binary definitions for each sleep measure: short sleep as ≤6 hours (compared with >6 hours), long sleep as ≥9 hours (compared with <9 hours), insomnia as WHIIRS≥10 (compared with WHIIRS<10) or WHIIRS≥6 if only 3 questions available (compared with WHIIRS<6), and EDS as ESS≥11 (compared with ESS<11). For OSA, we used REI (HCHS/SOL) and AHI (MESA and ARIC) to define moderate-to-severe OSA as REI or AHI≥15 (compared with REI or AHI<15).

#### Derivation of Global Cognitive Function Phenotypes

As illustrated in previous publications from the Cohorts for Heart and Aging Research in Genomic Epidemiology (CHARGE) consortium ^31,32^, global measures of cognitive function can be derived across cohorts based on a variety of cognitive assessments. Here, we extend this approach to longitudinal settings. Specifically, we derived a harmonized global cognitive function (GCF) measure across cohorts using a principal components analysis (PCA). First, we identified all cognitive tests available at baseline and follow-up visits in each cohort as described above. In each cohort separately, we then estimated sample means and standard deviations (SDs) for each test at baseline and derived standardized test scores at baseline and follow-up by subtracting the test-specific baseline mean and dividing by the baseline SD. Next, in each cohort separately, we performed PCA based on all baseline standardized test-specific scores available in the cohort. Baseline global cognitive function (GCF) was defined as the first PC (PC1) from this analysis. GCF at follow-up was then derived as the weighted summation of follow-up standardized test-specific scores available within each cohort, with weights being the baseline PCA loadings. In other words, both baseline and follow-up GCF are derived as weighted sums of baseline-standardized test scores with weights being baseline PC1 loadings. We used only individuals with complete cognitive data, i.e. available tests results for all cognitive tests used. Baseline and follow-up GCF variables were further standardized by subtracting the cohort-specific baseline mean and dividing by the cohort-specific baseline SD of PC1. Henceforth, we refer to these standardized measures as GCF (rather than standardized GCF) for simplicity. GCF change was defined as the GCF value at follow-up minus the GCF value at baseline. Finally, we defined “poor GCF” as age-adjusted PC1 > 1 SD below the median at each visit in each cohort. This cut-point is within the limits of most traditional definitions of mild cognitive impairment, and in line with previous studies which suggest a cut-point of 1 SD better balances sensitivity and specificity compared to other cut-points ^33^. Incident poor GCF was defined as not meeting this criterion at baseline but meeting this criterion at follow-up.

#### Statistical Analysis

Figure 1 provides an overview of study timelines and sample sizes (Panel A) and derivation of GCF measures (Panel B). After defining consensus sleep definitions and deriving GCF measures as previously described, we performed primary association analyses for the five binary sleep measures (short sleep, long sleep, insomnia, EDS, and OSA) and four cognitive outcomes (baseline GCF, GCF change, poor baseline GCF, and incident poor GCF) in each cohort. Linear regression was used for continuous outcomes (baseline GCF and GCF change) and logistic regression was used for binary outcomes (poor baseline GCF). For incident poor GCF, we used Poisson regression and included only individuals with healthy (not poor) GCF at baseline. Three adjustments sets were considered: the base model (M1) with age, sex, center, education level, *APOE4* (0, 1, or 2 copies), and race/ethnicity or Hispanic background (for HCHS/SOL); the cardiometabolic model (M2) with all variables from M1 plus hypertension (defined as >=140/90 or hypertension medication use), diabetes (defined as fasting glucose ≥126 mg/dL and fasting time >8 hours, fasting glucose ≥200 mg/dL and fasting time ≤8 hours, post-OGTT glucose ≥200 mg/dL, A1C ≥ 6.5%, or use of anti-diabetes medication), body mass index (BMI), high-density lipoprotein (HDL), low-density lipoprotein (LDL) and estimated glomerular filtration rate (eGFR); and a lifestyle model (M3) with all variables from M2 plus cigarette use and alcohol use. In addition, analyses of GCF change were also adjusted for time between baseline and follow-up visit and analyses of incident poor GCF included an offset term for the time between visits. Analyses in HCHS/SOL and MESA were also adjusted for language of test and analyses in HCHS/SOL accounted for its survey design (i.e. HCHS/SOL sampling weights in analyses of baseline cognitive measures and SOL-INCA sampling weights in analyses of follow-up cognitive measures). Cohort-specific estimates were then meta-analyzed using inverse-variance fixed-effects meta-analysis. Associations across the primary analyses (i.e. five sleep measures by four cognitive measures) with a Benjamini-Hochberg false-discovery rate (FDR) corrected p-value < 0.05 in meta-analyses were considered significant. Study-specific analyses were conducted using “glm” (for MESA and ARIC) and “surveyglm” (for HCHS/SOL) and meta-analyses were performed using “meta” in R.

**Figure 1.**
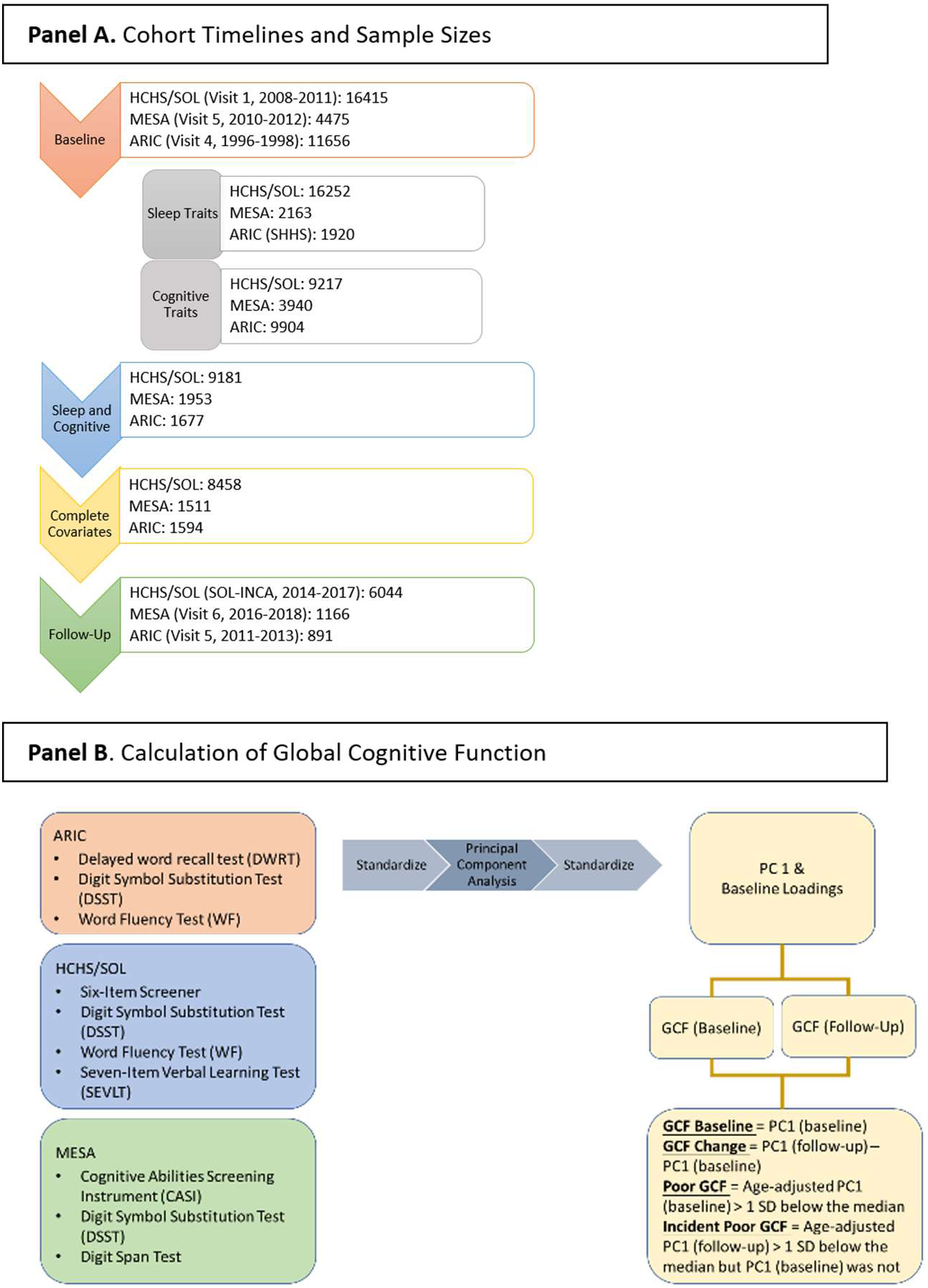
Study Framework. Sleep and cognitive measures were assessed at the following visits in each cohort, referred to as baseline as shown in Panel A: ARIC visit 4, HCHS/SOL visit 1, MESA visit 5. Cognitive measures were also assessed at follow-up visits in each cohort: ARIC visit 5, HCHS/SOL visit 2, MESA visit 6. Sample sizes for each cohort at baseline (sleep and cognition) and follow-up (cognition) are also shown. Global cognitive function was calculated in each cohort based on standardized test scores at baseline and follow-up, principal components analysis of baseline standardized test scores, and the projection of baseline PCs to follow-up as shown in Panel B. Baseline GCF defined as standardized PC1 at baseline, follow-up GCF as standardized PC1 at follow-up, GCF change as the GCF value at follow-up minus the GCF value at baseline, poor GCF as PC1 > 1 SD below the median, and incident poor GCF as not meeting this criterion at baseline but meeting this criterion at follow-up.

Since not all individuals who participated in baseline visits also participated in follow-up visits, the sample size was reduced between baseline (N=11,563) and follow-up (N=8,101). Because this could be a potential source of bias for analyses of change in GCF and incident poor GCF, we constructed inverse probability weights (IPW) to account for the probability of participating in follow-up visits based on baseline demographics (age, sex, race and ethnicity, study center, education, BMI, diabetes, hypertension, HDL, LDL, eGFR, alcohol and cigarette use) and ran sensitivity analyses implementing these weights. We also performed several other secondary analyses, including: (1) analyses of baseline GCF and change in GCF excluding individuals with poor GCF at baseline; (2) analyses of short sleep (≤6 hours) and long sleep (≥9 hours) compared to normal sleep (>6 hours and <9 hours) since some previous studies used this reference group to account for the U shape relationship between sleep duration and measures of cognitive function; (3) analyses with continuous sleep phenotypes as exposures, with WHIIRS harmonized by multiplying ARIC estimates by 12/20 to match the scale of HCHS/SOL and MESA estimates and AHI harmonized by scaling estimates in each cohort based on means and standard deviations; (4) sex-stratified analyses; and (5) analyses with individual cognitive tests, which were not always available or harmonizable across all cohorts though DSST from all three cohorts was harmonized by scaling estimates in each cohort based on means and standard deviations, B-SEVLT (Delayed Recall) from HCHS/SOL and DWRT from ARIC which are similar were harmonized by scaling estimates in each cohort based on means and standard deviations, and an SIS variable was created from subset of CASI questions in MESA to match the SIS from HCHS/SOL. Secondary analyses allowed us to explore whether associations that may be heterogeneous across cohorts are potentially more similar within certain strata and to address the robustness of our analytic approaches and potential specificity to cognitive domains. Secondary analyses used M1 covariates only.

## Results

### Cohorts Characteristics

Our analysis included 11,563 individuals at baseline (8,458 from HCHS/SOL, 1,511 from MESA and 1,594 from ARIC) and 8,101 individuals at follow-up (6,044 from HCHS/SOL, 1,166 from MESA and 891 from ARIC) (Figure 1 Panel A, Table 3, Supplemental Table 1). The average time between baseline and follow-up was 6.9 years in HCHS/SOL, 6.4 years in MESA, and 14.8 years in ARIC. Individuals from HCHS/SOL tended to be younger at baseline (average age 55.2 years) than individuals from MESA (68.6 years) and ARIC (62.8 years). HCHS/SOL also included more females (62.0%) than MESA (53.9%) and ARIC (51.0%). Education level varied across cohorts with 36.8% of participants in HCHS/SOL, 69.0% in MESA and 50.9% in ARIC attending school beyond high school. Other factors including mean levels of BMI, LDL and HDL were similar across cohorts.

**Table 3.** Study Characteristics among Individuals with Sleep Phenotype and Global Cognitive Function Data.

| Characteristics (Continuous) | HCHS/SOL<sup>a</sup> |  |  | MESA |  |  | ARIC |  |  |
| --- | --- | --- | --- | --- | --- | --- | --- | --- | --- |
|  | N | Mean | SD | N | Mean | SD | N | Mean | SD |
| Age at Baseline Visit (years)<sup>b</sup> | 8458 | 55.2 | 7.4 | 1511 | 68.6 | 9.2 | 1594 | 62.8 | 5.6 |
| Age at Follow-Up Visit (years)<sup>c</sup> | 6044 | 62.0 | 7.2 | 1166 | 73.7 | 8.6 | 891 | 75.8 | 5.1 |
| Years between Baseline and Follow-up Visit | 6044 | 6.9 | 1.2 | 1166 | 6.4 | 0.6 | 891 | 14.8 | 1.0 |
| Global Cognitive Function (PC1) at Baseline Visit | 8458 | 0.01 | 1.0 | 1511 | 0.01 | 1.0 | 1594 | 0.2 | 1.2 |
| Global Cognitive Function (PC1) at Follow-Up Visit | 5949 | 0.02 | 1.0 | 718 | 0.03 | 0.9 | 891 | 0.2 | 1.3 |
| Sleep Duration (hours) | 8137 | 7.8 | 1.4 | 1492 | 8.0 | 1.4 | 1555 | 7.4 | 1.0 |
| Insomnia WHIIRS | 8272 | 7.7 | 5.5 | 1491 | 7.5 | 4.5 | 1582 | 4.9 | 2.4 |
| Daytime Sleepiness ESS | 8363 | 6.0 | 5.0 | 1496 | 6.2 | 4.1 | 1556 | 8.3 | 4.3 |
| REI / AHI | 7605 | 8.1 | 12.8 | 1390 | 20.0 | 18.7 | 1430 | 9.6 | 12.7 |
| BMI (kg/m<sup>2</sup>) | 8438 | 30.1 | 5.7 | 1510 | 29.1 | 5.5 | 1592 | 28.7 | 5.0 |
| HDL (mg/dL) | 8433 | 50.0 | 13.3 | 1496 | 55.8 | 16.6 | 1594 | 49.2 | 16.0 |
| LDL (mg/dL) | 8248 | 129.4 | 37.5 | 1488 | 106.5 | 32.0 | 1571 | 122.3 | 32.4 |
| Characteristics (Categorical) | N | % |  | N | % |  | N | % |  |
| **Sex** |  |  |  |  |  |  |  |  |  |
| Male | 3218 | 38.0 |  | 696 | 46.1 |  | 781 | 49.0 |  |
| Female | 5240 | 62.0 |  | 815 | 53.9 |  | 813 | 51.0 |  |
| **Short Sleep (≤6 Hours)** |  |  |  |  |  |  |  |  |  |
| No | 7249 | 85.7 |  | 1364 | 90.3 |  | 1426 | 89.5 |  |
| Yes | 888 | 10.5 |  | 128 | 8.5 |  | 129 | 8.1 |  |
| Missing | 321 | 3.8 |  | 19 | 1.3 |  | 39 | 2.4 |  |
| **Long Sleep (≥9 Hours )** |  |  |  |  |  |  |  |  |  |
| No | 6451 | 76.3 |  | 1136 | 75.2 |  | 1466 | 92.0 |  |
| Yes | 1686 | 19.9 |  | 356 | 23.6 |  | 89 | 5.6 |  |
| Missing | 321 | 3.8 |  | 19 | 1.3 |  | 39 | 2.4 |  |
| **Insomnia (WHIIRS≥10 or ≥6 if 3 Ques)** |  |  |  |  |  |  |  |  |  |
| No | 5266 | 62.3 |  | 1034 | 68.4 |  | 755 | 47.4 |  |
| Yes | 3006 | 35.5 |  | 457 | 30.2 |  | 827 | 51.9 |  |
| Missing | 188 | 2.2 |  | 20 | 1.3 |  | 12 | 0.8 |  |
| **Excessive Daytime Sleepiness (ESS≥11)** |  |  |  |  |  |  |  |  |  |
| No | 6958 | 82.3 |  | 1285 | 85.0 |  | 1112 | 69.8 |  |
| Yes | 1405 | 16.6 |  | 211 | 14.0 |  | 444 | 27.9 |  |
| Missing | 95 | 1.1 |  | 15 | 1.0 |  | 38 | 2.4 |  |
| **OSA (REI / AHI ≥ 15 )** |  |  |  |  |  |  |  |  |  |
| No | 6425 | 76.0 |  | 723 | 47.9 |  | 1143 | 71.7 |  |
| Yes | 1180 | 14.0 |  | 667 | 44.2 |  | 287 | 18.0 |  |
| Missing | 853 | 10.1 |  | 121 | 8.0 |  | 164 | 10.3 |  |
| **Education** |  |  |  |  |  |  |  |  |  |
| No High School Diploma | 3502 | 41.4 |  | 210 | 13.9 |  | 172 | 10.8 |  |
| At Most a High School Diploma | 1841 | 21.8 |  | 258 | 17.1 |  | 611 | 38.3 |  |
| Greater than High School Education | 3115 | 36.8 |  | 1043 | 69.0 |  | 811 | 50.9 |  |
| **APOE4** |  |  |  |  |  |  |  |  |  |
| 0 | 6589 | 77.9 |  | 1088 | 72.0 |  | 1126 | 70.6 |  |
| 1 | 1743 | 20.6 |  | 390 | 25.8 |  | 422 | 26.5 |  |
| 2 | 126 | 1.5 |  | 33 | 2.2 |  | 46 | 2.9 |  |

a) Means, standard deviations (SD), and percentages (%) for HCHS/SOL presented in this table are not weighted for the study sampling weights. For weighted means, standard deviations (SD), and percentages (%) see Supplemental Table 1.
b) Baseline was HCHS/SOL visit 1, MESA visit 5, and ARIC visit 4
c) Follow-up was HCHS/SOL visit 2, MESA visit 6, and ARIC visit 5

### Global cognitive function measures

Measures of GCF in HCHS/SOL, MESA, and ARIC were based on the first PC derived from PC analysis of individual cognitive tests (PC loadings are presented in Supplemental Figure 1). The first PC (PC1) explained 52-56% of variance in each study, compared to the second or later PCs which each explained <17-27% of variance depending on the study (Supplemental Figure 2). Moreover, the GCF measure defined by PC1 was highly correlated with individual cognitive assessments in each study: r^2^ ranged from 0.54-0.84 in HCHS/SOL, 0.68-0.78 in MESA, and 0.69-0.82 in ARIC depending on the test (Supplemental Figure 3).

### Sleep Phenotypes and Baseline Cognitive Function

In meta-analyses of baseline GCF, long sleep was significantly associated with lower GCF at baseline (≥9 hours, β=-0.19, SE=0.03, p=3.78x10^-14^, FDR p=7.56x10^-13^, p for heterogeneity=0.78, Figure 2, Supplemental Table 2). Insomnia (WHIIRS ≥10 or 3 questions ≥6 for ARIC) was also suggestively associated with lower baseline GCF (β=-0.051, SE=0.02, p=0.01, FDR p=0.07, p for heterogeneity=0.005). When baseline GCF was dichotomized such that age-adjusted values lower than 1 SD below the median were considered “poor” GCF, both long sleep (OR=1.77, 95% CI=1.49-2.09, p=2.96x10^-11^, FDR p=2.96x10^-10^, p for heterogeneity=0.22) and insomnia (OR=1.22, 95% CI=1.06-1.41, p=6.76x10^-3^, FDR p=0.05, p for heterogeneity=0.01) were associated with increased risk of poor baseline GCF (Figure 2, Supplemental Table 2). In contrast, EDS, short sleep and OSA were not associated with continuous baseline GCF nor dichotomized poor baseline GCF in meta-analyses.

**Figure 2.**
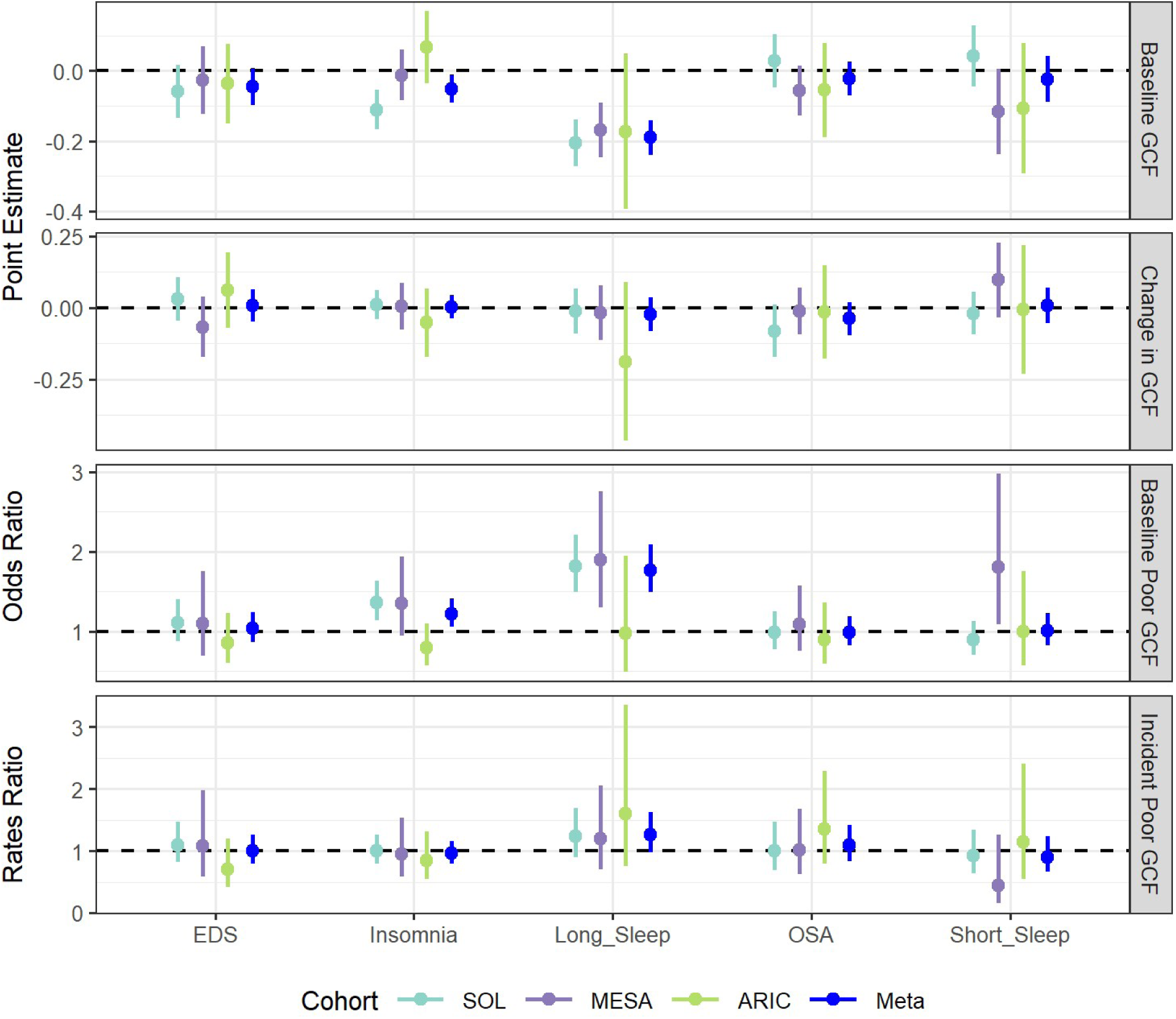
Study-Specific and Meta-Analysis Results (Estimates and 95% Confidence Intervals) for the Association between Sleep Phenotypes and Global Cognitive Function Measures. Estimates adjusted for age, sex, center, education level, *APOE4*, race/ethnicity or Hispanic background (for HCHS/SOL), and language of test (HCHS/SOL and MESA). Analyses in HCHS/SOL accounted for its survey design. EDS= Excessive Daytime Sleepiness, OSA=Obstructive Sleep Apnea, GCF=Global Cognitive Function.

Long sleep continued to be associated with lower baseline GCF when individuals with poor baseline GCF were excluded (β=-0.11, SE=0.02, p=1.32x10^-6^, p for heterogeneity=0.57, Supplemental Table 3). Further, long sleep was associated with baseline GCF (β=-0.19, SE=0.03, p=1.21x10^-14^, p for heterogeneity=0.60) and poor GCF (OR=1.87, 95% CI=1.57-2.22, p=1.62x10^-12^, p for heterogeneity=0.55) when using normal sleepers (>6 and <9 hours) as the reference group (Supplemental Table 4). In contrast, insomnia was no longer associated with baseline GCF when excluding individuals with poor GCF at baseline (β=-0.02, SE=0.02, p=0.20, p for heterogeneity=0.11, Supplemental Table 3). Interestingly, the association between EDS and lower baseline GCF was strengthened when excluding individuals with poor baseline GCF (β=-0.05, SE=0.02, p=0.02, p for heterogeneity=0.80, Supplemental Table 3) and the associations between short sleep and lower baseline GCF (β=-0.07, SE=0.03, p=0.04, p for heterogeneity=0.14, Supplemental Table 4) and poor GCF (OR=1.22, 95% CI=0.99-1.50, p=0.05, p for heterogeneity=0.05, Supplemental Table 4) were strengthened when compared to normal sleepers. When we considered continuous measures of sleep, sleep duration was associated with baseline GCF (β=-0.034, SE=0.007, p=6.74x10^-7^, p for heterogeneity=0.09) and poor GCF (OR=1.12, 95% CI=1.07-1.18, p=1.12x10^-5^, p for heterogeneity=0.37) (Supplemental Table 5,

Supplemental Figure 4). WHIIRS score was also associated with lower baseline GCF (β=-0.005, SE=0.002, p=3.49x10^-3^, p for heterogeneity=0.02) and poor GCF (OR=1.02, 95% CI=1.01-1.04, p=8.26x10^-4^, p for heterogeneity=0.47) in meta-analyses. Continuous measures of ESS and REI/AHI levels were not associated with continuous baseline GCF nor dichotomized poor baseline GCF in meta-analyses.

The results just described were for the primary adjustment set (age, sex, center, education level, *APOE4* and race/ethnicity). Associations were similar across other adjustment sets. In particular, associations of long sleep and insomnia with baseline GCF and poor GCF remained after adjustment for hypertension, diabetes, BMI, HDL, LDL, and eGFR (Supplemental Table 6) as well as cigarette use and alcohol use (Supplemental Table 7). In secondary sex-stratified analyses, results were generally similar among males and females (Supplemental Figure 5).

Finally, we noted several associations between sleep measures and individual cognitive assessments at baseline (Figure 3, Supplemental Table 8). In particular, long sleep was significantly associated with lower scores at baseline for the following assessments: DSST (β=-0.17, SE=0.02, p=4.83x10^-14^, FDR p=2.90x10^-12^, p for heterogeneity=0.37 available in all 3 cohorts); SIS (β=-0.16, SE=0.03, p=8.17x10^-10^, FDR p=2.45x10^-8^, p for heterogeneity=0.75 available in HCHS/SOL and MESA); B-SEVLT Delayed Recall or DWRT (β=-0.14, SE=0.03, p=9.17x10^-6^, FDR p=1.38x10^-4^, p for heterogeneity=0.44 available in HCHS/SOL and ARIC); WFT (β=-0.13, SE=0.04, p=8.79x10^-4^, FDR p=0.009, p for heterogeneity=0.83 available in HCHS/SOL and ARIC); and B-SEVLT Sum (β=-0.13, SE=0.03, p=1.29x10^-4^, FDR p=1.55x10^-3^ available in HCHS/SOL). Likewise, insomnia was significantly associated with lower scores at baseline for DSST (β=-0.05, SE=0.02, p=0.006, FDR p=0.04, p for heterogeneity=0.002 available in all 3 cohorts); B-SEVLT Delayed Recall or DWRT (β=-0.07, SE=0.02, p=0.004, FDR p=0.03, p for heterogeneity=0.24 available in HCHS/SOL and ARIC); and B-SEVLT Sum (β=-0.13, SE=0.03, p=3.12x10^-6^, FDR p=6.25x10^-5^ available in HCHS/SOL). EDS, OSA and short sleep were not associated with any of the individual cognitive assessments at baseline.

**Figure 3.**
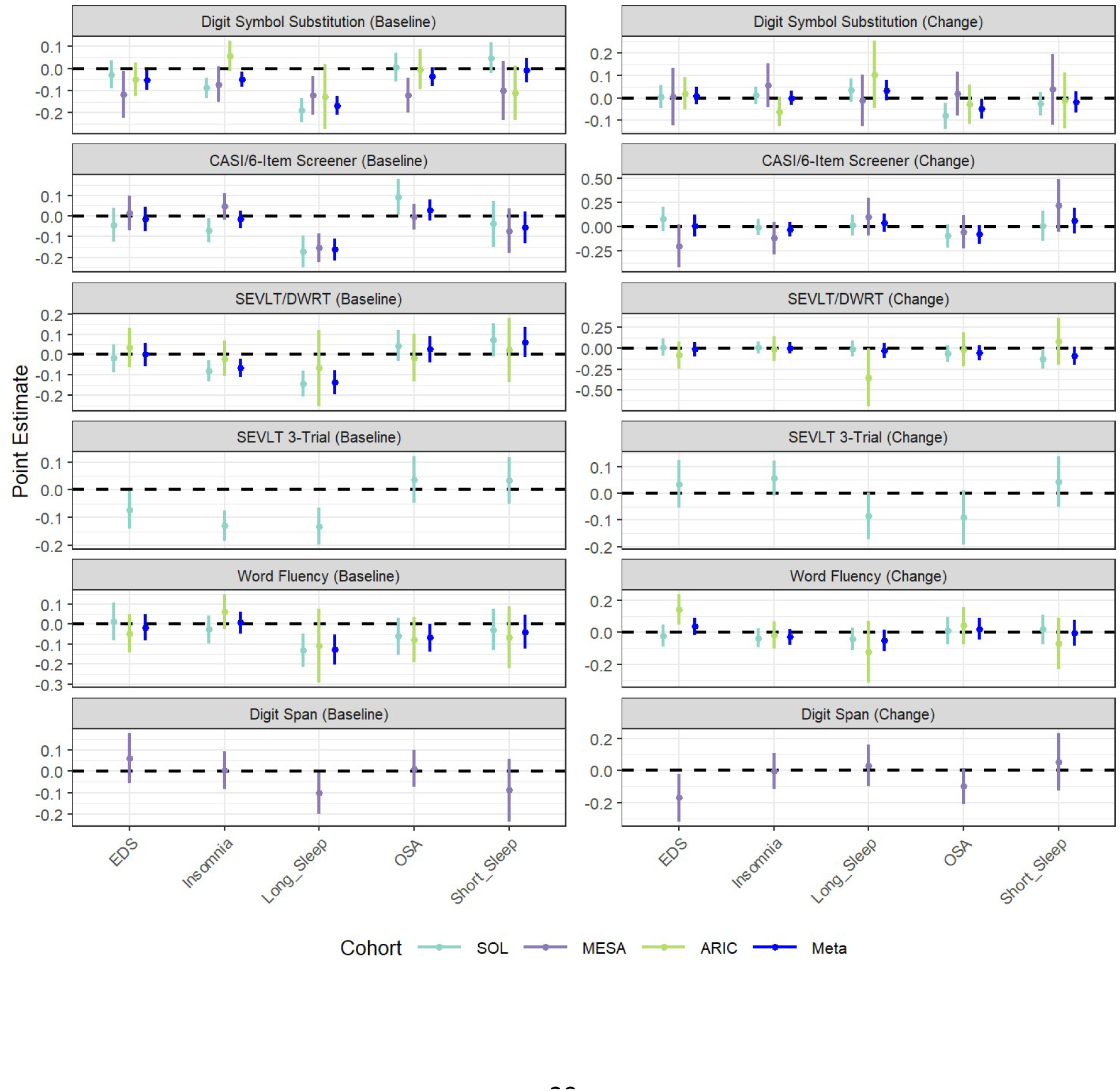
Study-Specific and Meta-Analysis Results (Estimates and 95% Confidence Intervals) for the Association between Sleep Phenotypes and Single Cognitive Assessments. Estimates adjusted for age, sex, center, education level, *APOE4*, race/ethnicity or Hispanic background (for HCHS/SOL), and language of test (HCHS/SOL and MESA). Analyses in HCHS/SOL accounted for its survey design. Single cognitive assessments were not always available or harmonizable across all cohorts though DSST from all three cohorts was harmonized by scaling estimates in each cohort based on means and standard deviations, B-SEVLT (Delayed Recall) from HCHS/SOL and DWRT from ARIC which are similar were harmonized by scaling estimates in each cohort based on means and standard deviations, and an SIS variable was created from subset of CASI questions in MESA to match the SIS from HCHS/SOL. EDS= Excessive Daytime Sleepiness, OSA=Obstructive Sleep Apnea, GCF=Global Cognitive Function, CASI=Cognitive Abilities Screening Instrument, SEVLT= Spanish English Verbal Learning Test, DWRT=Delayed Word Recall Test.

### Sleep Phenotypes and Change in Cognitive Function from Baseline to Follow-up

Sleep measures (binary or continuous) were not associated with change in GCF from baseline to follow-up nor with incident poor cognitive function (Figure 2, Supplemental Table 2, Supplemental Table 5, Supplemental Figure 4). The strongest, though not significant, association was long sleep with a greater likelihood of incident poor GCF (OR=1.26, 95%CI=0.98-1.63, p-value=0.08, FDR p=0.30, p for heterogeneity=0.80). When compared to normal sleepers, the association between long sleep and incident poor GCF was similar (OR=1.28, 95% CI=0.99-1.66, p-value=0.06, p for heterogeneity=0.50, Supplemental Table 4). Since the number of individuals participating in the follow-up visit was reduced compared to the baseline visit across all cohorts, we also ran sensitivity analyses of change in GCF and incident poor GCF using IPW to account for the probability of participating in follow-up visits. Reassuringly, results were similar without and with IPW (Supplemental Table 9, Supplemental Figure 6). In particular, long sleep remained suggestively associated with greater risk of incident poor GCF (OR=1.23, 95%CI=0.96-1.58, p-value=0.11, p for heterogeneity=0.34). Estimates were also similar when additionally adjusted for hypertension, diabetes, BMI, HDL, LDL, and eGFR (Supplemental Table 6) as well as cigarette use and alcohol use (Supplemental Table 7). In sex-stratified analyses, most associations were similar among males and females (Supplemental Figure 5), though long sleep was associated with increased odds of incident poor GCF in males (OR=1.50, 95% CI=1.02-2.20, p=0.04) but not females (OR=1.13, 95%CI=0.83-1.55, p=0.44). Finally, none of the sleep measures appeared to be associated with changes in scores for individual cognitive assessments (Figure 3, Supplemental Table 8).

## Discussion

In this large meta-analysis, we found consistent evidence that long sleep was associated with global cognitive function measures harmonized across three studies. Specifically, long sleep was associated with lower GCF at baseline and higher likelihood of poor GCF at baseline. Associations between long sleep and lower baseline GCF remained strong even when excluding individuals with poor baseline GCF or when using normal sleepers as the referent. With respect to more specific domains of cognitive function, long sleep was associated with lower baseline measures of executive function and information processing (DSST), episodic memory (B-SEVLT or DWRT) and language (WFT). Insomnia symptoms were also suggestively associated with lower GCF at baseline and higher likelihood of poor GCF at baseline, including lower baseline measures of executive function and information processing (DSST) and episodic memory (B-SEVLT or DWRT). However, the association between insomnia and lower baseline GCF was weaker when excluding individuals with poor baseline GCF.

Previous studies of sleep duration and cognitive function have reported mixed results. In a pooled analysis of two prospective studies (English Longitudinal Study of Ageing and China Health and Retirement Longitudinal Study) including over 28,000 individuals, both short sleep (defined there as ≤4 hours) and long sleep (defined there as ≥10 hours) were significantly associated with faster decline in global cognitive z scores based on several tests when compared with individuals in the referent group (7 hours) ^34^. Likewise, in a large meta-analysis of 11 cross-sectional and 7 prospective cohort studies encompassing over 97,000 individuals, self-reported short sleep (OR=1.40, 95% CI=1.27-2.56) and long sleep (OR=1.58, 95% CI=1.43-1.74) were associated with increased odds of poor cognitive function ^6^. However, this meta-analysis noted significant heterogeneity among study estimates possibly due to heterogeneity in exposure-outcome associations by population as well as the variety of sleep definitions (e.g., short sleep varied between <4 to <7 hours and long sleep thresholds between >7 to ≥11 hours, with the most common referent group defined as 7 hours or 6-8 hours of sleep) and cognitive assessments (e.g., single tests vs. global cognitive measures calculated across many tests) utilized by studies. Among the 11 cross-sectional studies included in the meta-analysis, four studies reported associations for both short and long sleep ^35–38^, four studies reported associations only for long sleep ^39–42^, two studies reported associations only for short sleep ^43,44^, and one study reported no associations ^45^ with prevalent cognitive measures. Among the seven prospective studies included in this meta-analysis, three studies reported associations for both short and long sleep ^46–48^, two studies reported associations only for long sleep ^49,50^, and two studies reported associations only for short sleep ^51,52^ with incident cognitive measures. In a smaller meta-analysis of three case-control studies and seven cohort studies examining only long sleep, associations with cognitive decline (RR=1.42, 95% CI=1.27-1.59) and cognitive impairment (RR=1.38, 95% CI=1.23-1.56) were reported ^8^. Recent studies not included in previous meta-analyses have also reported a variety of results^53–56^.

In our study, we defined short and long sleep as ≤6 hours and ≥9 hours, respectively. These thresholds are similar to several previous studies and based on our previous work ^30^. To maximize sample size, our main analyses compared ≤6hours with >6hours and ≥9hours with <9hours. Since many of the previous studies considered normal sleepers as the referent group, and due to the U-shaped association between sleep duration and cognitive function, we also performed sensitivity analyses contrasting short sleepers (≤6 hours) and long sleepers (≥9 hours) with normal sleepers (>6 to <9 hours). For long sleep, associations with baseline GCF and baseline poor GCF were consistent across studies and similar in meta-analyses using either referent group. This may be due to the large number of individuals classified as long sleepers in HCHS/SOL (>20% of individuals were long sleepers in HCHS/SOL and MESA compared to 6% in ARIC), coupled with the strong association of long sleep with GCF. For short sleep, associations with baseline GCF and baseline poor GCF were not significant in the main analysis (referent group >6 hours), but were stronger in the sensitivity analyses (referent group >6 to <9 hours).

Results for the association between short sleep and baseline GCF in HCHS/SOL appeared to differ from those in MESA and ARIC in the main analysis, but less so in the sensitivity analysis. Moreover, the short sleep with baseline GCF results for MESA were strengthened using normal sleepers as the referent, which may have been a driving factor for the overall association strength in the secondary meta-analyses.

For insomnia, two previous meta-analyses have systematically considered associations with cognition measures, reporting mixed results ^3,4,57,58^. One meta-analysis included 24 studies covering 1,197 individuals (639 with insomnia and 558 normal sleepers), finding general cognitive function did not significantly differ between those with and without insomnia (effect=-0.12, 95%CI: -0.42 to 0.18) ^3^. For specific cognitive domains, lower performance among those with insomnia was noted for episodic memory, working memory, and problem solving, but none of the other domains considered. In contrast, a second larger meta-analysis which included 48 studies covering 4,539 individuals, reported insomnia was associated with lower overall cognitive performance (effect=-0.19, 95%CI: -0.38 to 0.00) ^4^. However, associations again varied by domain. Those with insomnia symptoms performed worse than normal sleepers with respect to working memory, episodic memory, and problem solving as well as perceptual processes, alertness, and complex reaction time, but not the other domains considered. Both meta-analyses noted significant heterogeneity across study effects, with insomnia in previous studies using varying information from self-report, PSG, and clinical criteria based on International Classification of Disease (ICD) or Diagnostic and Statistical Manual of Mental Disorders (DSM) and cognitive assessments varying between different domain-specific tools and global measures ^3,57^. Although several previous studies defined insomnia based on ICD or DSM codes, these metrics are not widely available in prospective cohort studies. Furthermore, insomnia is frequently undiagnosed ^59^. Therefore, using ICD or DSM codes to define insomnia can lead to misclassification.

In our study we used the validated WHIIRS scale to classify individuals according to insomnia symptoms. Although one of the cohorts in our study asked three of the five standard WHIIRS questions while the other two studies asked all five questions, previous research showed that a polygenic risk score (PRS) for insomnia was similarly and strongly associated with both WHIIRS 5 questions ≥10 and WHIIRS 3 questions ≥6 ^30^. We found that insomnia was associated with both baseline GCF and baseline poor GCF, though significant heterogeneity was noted for both (p for heterogeneity < 0.05). Among cohort-specific estimates, insomnia was significantly associated with lower baseline GCF in HCHS/SOL (β=-0.11, SE=0.03. p=1.35x10^-4^), but estimates in ARIC (β=0.06, SE=0.05, p=0.23) and MESA (β=-0.007, SE=0.04, p=0.86) were not significant and closer to null. Therefore, the findings for insomnia may largely be driven by HCHS/SOL. Cohort differences may reflect differences in social factors across cohorts that may contribute to insomnia symptoms.

Interestingly, neither OSA nor EDS were not associated with GCF measures in our study. OSA obtained from overnight sleep studies was defined as AHI or REI ≥15 and EDS was defined using the validated ESS with a threshold ≥11 across the cohorts in our meta-analysis. Although a previous meta-analysis of 19 studies reported evidence of an overall association for OSA with cognitive decline, significant heterogeneity across study-specific estimates was noted likely due to the wide range of AHI thresholds (>5 to ≥30) and variety of cognitive domains assessed ^5^. In addition, a review of over 60 articles regarding cognitive function and OSA (ranging from mild/moderate to severe) and EDS (usually defined by ESS score), noted cognitive decline may be limited to OSA patients with EDS, and not as apparent in individuals without EDS ^60^. This review also noted variation of findings, though, depending on the cognitive assessments and domains considered, as well as potential modification by CPAP utilization. While associations could be influenced by OSA treatment, in our study individuals using CPAP at baseline were excluded and over time CPAP use was very low.

With respect to cognitive measures, many previous studies relied on a single test to define outcomes. In our study, we calculated global cognitive function based on several cognitive tests, similar to the approach used in previous CHARGE GWAS ^31,32^. Since cohorts often utilize a wide array of tests that are not uniform across studies, global measures provide comprehensive and standardized metrics that facilitate comparison and aggregation across studies. To better understand which domains of cognition were particularly impacted by sleep, though, we additionally considered associations with individual tests. Similar to previous studies ^3,4,6^, we noted long sleep and insomnia were associated with executive function and information processing (DSST) and episodic memory (B-SEVLT or DWRT). In addition, previous studies employed a variety of study designs including both cross-sectional and prospective cohort. By utilizing the prospective nature of the cohorts in our study, we considered both prevalent and longitudinal cognitive measures. Although we found strong associations between long sleep and insomnia with baseline cognitive measures, sleep measures were not associated with change in cognitive measures in our study. Cohorts in our study had a mean age at baseline and mean follow-up times as follows: 62.8 (14.8) years in ARIC, 68.6 (6.3) years in MESA and 55.2 (6.9) years in HCHS/SOL. Therefore, it is possible that we found more associations with baseline measures than follow-up measures because older participants may have already experienced some cognitive impairment prior to or at baseline and longer follow-up times are needed to capture further decline. Further, since cognitive measures were available at a single follow-up visit, estimates for change in cognitive measures may have been noisier and weaker than if we had additional follow-up visits to establish more stable trends.

To address these potential limitations, we performed analyses of baseline GCF and change in GCF excluding individuals with poor baseline GCF (defined as 1 SD below the median based on previous studies suggesting this threshold ^33^) and analyses of change in GCF and incident poor GCF using IPW. For baseline GCF, we continued to note strong associations for long sleep but weaker associations for insomnia when excluding individuals with poor baseline GCF. For change in GCF, results excluding individuals with poor baseline GCF and results based on IPW were similar to results from the main analyses. Despite our attempt to account for selection bias due to death and visit nonattendance through IPW, it is possible that some bias remained, with participants who experienced greater cognitive impairment less likely to attend follow-up visits.

Another potential limitation is that sleep measures other than OSA were based on self-reported data. However, we utilized thresholds of sleep measures within meaningful limits based on previous research to help mitigate misclassification. Further, a study comparing self-reported sleep data (i.e. duration) with actigraphy data in HCHS/SOL previously showed moderate correlation ^64^. Since sleep measures were not available at follow-up, we were unable to consider how sleep measures may have changed as cognitive function or life circumstances changed (e.g., some participants may have transitioned from working to retirement, with changes in sleep habits). In particular, if individuals with cognitive impairment experience an onset of sleep disturbances this could potentially lead to reverse causation. Further, individuals with comorbidities may also experience more sleep disturbances. While additional adjustment for hypertension, diabetes, BMI, HDL, LDL and eGFR did not materially change sleep and cognition estimates in our study, we did not have uniform information on other health conditions, such as depression, to consider. As the cohorts in our meta-analysis continue to collect prospective data it will be informative to further track and analyze changes in sleep patterns and cognitive measures.

In summary, our study was strengthened by and extended previous literature by uniformly defining sleep and cognitive measures across three large, diverse cohorts representing different samples of the U.S. population. We found consistent evidence that self-reported long sleep was associated with lower baseline GCF and increased odds of poor baseline GCF across cohorts regardless of referent group or exclusions based on GCF. Insomnia was also suggestively associated with lower baseline GCF and increased odds of poor baseline GCF, though results varied slightly by study with the strongest associations in HCHS/SOL and associations with baseline GCF were weaker when excluding individuals with poor baseline GCF. Our meta-analysis highlights the important relationship between specific domains of sleep and cognitive function in older adults. Additional prospective data is needed to further elucidate associations with change in cognitive function across time.

## Data and code availability statement

In accordance with participants informed consent, HCHS/SOL and MESA data are available through data use agreement in dbGaP according to the study specific accessions: HCHS/SOL in phs000810 and MESA in phs000209. HCHS/SOL data also are available via data use agreement with the HCHS/SOL Data Coordinating Center (DCC) at the University of North Carolina at Chapel Hill, see collaborators website: https://sites.cscc.unc.edu/hchs/. Researchers can email the HCHS/SOL DCC at. MESA data are also available from BioLINCC (https://biolincc.nhlbi.nih.gov/studies/mesa/) or through application to the MESA Coordinating Center. Data from the ARIC study are available upon request to the ARIC Data Coordinating Center as described on the ARIC study website (<u>Obtain/Submit Data | ARIC</u>). Example analysis code available on GitHub: https://github.com/YuzBioStat/Sleep_Cog_Association

## Acknowledgements

The research presented in this paper was supported by US Department of Health and Human Services, National Institutes of Health, National Institute of Aging (R01AG080598) and National Heart, Lung, and Blood Institute (R01HL161012).

The Atherosclerosis Risk in Communities study has been funded in whole or in part with Federal funds from the National Heart, Lung, and Blood Institute, National Institutes of Health, Department of Health and Human Services, under Contract nos. (75N92022D00001, 75N92022D00002, 75N92022D00003, 75N92022D00004, 75N92022D00005). The authors thank the staff and participants of the ARIC study for their important contributions. The ARIC Neurocognitive Study is supported by U01HL096812, U01HL096814, U01HL096899, U01HL096902, and U01HL096917 from the NIH (NHLBI, NINDS, NIA and NIDCD).

The Hispanic Community Health Study/Study of Latinos is a collaborative study supported by contracts from the National Heart, Lung, and Blood Institute (NHLBI) to the University of North Carolina (75N92025D00001, HHSN268201300001I / N01-HC-65233), University of Miami (75N92025D00009, HHSN268201300004I / N01-HC-65234), Albert Einstein College of Medicine (75N92025D00007, HHSN268201300002I / N01-HC-65235), University of Illinois at Chicago (75N92025D00008, HHSN268201300003I / N01-HC-65236 Northwestern Univ), and San Diego State University (75N92025D00010, HHSN268201300005I / N01-HC-65237). The following Institutes/Centers/Offices have contributed to the HCHS/SOL through a transfer of funds to the NHLBI: National Institute on Minority Health and Health Disparities, National Institute on Deafness and Other Communication Disorders, National Institute of Dental and Craniofacial Research, National Institute of Diabetes and Digestive and Kidney Diseases, National Institute of Neurological Disorders and Stroke, NIH Institution-Office of Dietary Supplements.

Support for MESA is provided by contracts 75N92025D00022, 75N92020D00001, HHSN268201500003I, N01-HC-95159, 75N92025D00026, 75N92020D00005, N01-HC-95160, 75N92020D00002, N01-HC-95161, 75N92025D00024, 75N92020D00003, N01-HC-95162, 75N92025D00027, 75N92020D00006, N01-HC-95163, 75N92025D00025, 75N92020D00004, N01-HC-95164, 75N92025D00028, 75N92020D00007, N01-HC-95165, N01-HC-95166, N01-HC-95167, N01-HC-95168, N01-HC-95169, UL1-TR-000040, UL1-TR-001079, UL1-TR-001420, UL1TR001881, DK063491, and R01HL105756. The MESA Sleep Ancillary studies were funded by NIH-NHLBI R01HL098433 and NIH-NIA 5R01AG070867. The authors thank the MESA participants and the MESA investigators and staff for their valuable contributions. A full list of participating MESA investigators and institutions can be found at http://www.mesa-nhlbi.org.

