## Supplemental for "Long Sleep and Insomnia are Associated with Reduced Global Cognitive Function across Diverse Populations"

**Supplemental Table 1. Study Characteristics among Individuals with Sleep Phenotype and Global Cognitive Function Data, With Weighted Values for HCHS/SOL**

| | HCHS/SOL[a] | | | MESA | | | ARIC | | |
|---|---|---|---|---|---|---|---|---|---|
| **Characteristics (Continuous)** | **N** | **Mean** | **SD** | **N** | **Mean** | **SD** | **N** | **Mean** | **SD** |
| Age at Baseline Visit (years) | 8458 | 56.2 | 8.1 | 1511 | 68.6 | 9.2 | 1594 | 62.8 | 5.6 |
| Age at Follow-Up Visit (years)[c] | 6044 | 63.0 | 7.9 | 1166 | 73.7 | 8.6 | 891 | 75.8 | 5.1 |
| Years between Baseline and Follow-up Visit | 6044 | 7.0 | 1.2 | 1166 | 6.4 | 0.6 | 891 | 14.8 | 1.0 |
| Global Cognitive Function (PC1) at Baseline Visit | 8458 | -0.03 | 1.0 | 1511 | 0.01 | 1.0 | 1594 | 0.2 | 1.2 |
| Global Cognitive Function (PC1) at Follow-Up Visit | 5949 | -0.02 | 1.0 | 718 | 0.03 | 0.9 | 891 | 0.2 | 1.3 |
| Sleep Duration (hours) | 8137 | 7.8 | 1.4 | 1492 | 8.0 | 1.4 | 1555 | 7.4 | 1.0 |
| Insomnia WHIIRS | 8272 | 7.7 | 5.6 | 1491 | 7.5 | 4.5 | 1582 | 4.9 | 2.4 |
| Daytime Sleepiness ESS | 8363 | 5.9 | 5.0 | 1496 | 6.2 | 4.1 | 1556 | 8.3 | 4.3 |
| REI / AHI | 7605 | 8.8 | 13.4 | 1390 | 20.0 | 18.7 | 1430 | 9.6 | 12.7 |
| BMI (kg/m$^2$) | 8438 | 29.8 | 5.5 | 1510 | 29.1 | 5.5 | 1592 | 28.7 | 5.0 |
| HDL (mg/dL) | 8433 | 49.7 | 13.3 | 1496 | 55.8 | 16.6 | 1594 | 49.2 | 16.0 |
| LDL (mg/dL) | 8248 | 129.8 | 38.2 | 1488 | 106.5 | 32.0 | 1571 | 122.3 | 32.4 |
| **Characteristics (Categorical)** | **N** | **%** | | **N** | **%** | | **N** | **%** | |
| **Sex** | | | | | | | | | |
| Male | 2586.1 | 45.4 | | 696 | 46.1 | | 781 | 49.0 | |
| Female | 3115.5 | 54.6 | | 815 | 53.9 | | 813 | 51.0 | |
| **Short Sleep (≤6 Hours)** | | | | | | | | | |
| No | 4885.7 | 89.4 | | 1364 | 90.3 | | 1426 | 89.5 | |
| Yes | 578.3 | 10.6 | | 128 | 8.5 | | 129 | 8.1 | |
| Missing | | | | 19 | 1.3 | | 39 | 2.4 | |
| **Long Sleep (≥9 Hours )** | | | | | | | | | |
| No | 4295.1 | 78.6 | | 1136 | 75.2 | | 1466 | 92.0 | |
| Yes | 1168.9 | 21.4 | | 356 | 23.6 | | 89 | 5.6 | |
| Missing | | | | 19 | 1.3 | | 39 | 2.4 | |
| **Insomnia (WHIIRS≥10 or ≥6 if 3 Ques)** | | | | | | | | | |
| No | 3571.6 | 64.2 | | 1034 | 68.4 | | 755 | 47.4 | |
| Yes | 1989.7 | 35.8 | | 457 | 30.2 | | 827 | 51.9 | |
| Missing | | | | 20 | 1.3 | | 12 | 0.8 | |
| **Excessive Daytime Sleepiness (ESS≥11 )** | | | | | | | | | |
| No | 4683.0 | 83.2 | | 1285 | 85.0 | | 1112 | 69.8 | |
| Yes | 943.4 | 16.8 | | 211 | 14.0 | | 444 | 27.9 | |
| Missing | | | | 15 | 1.0 | | 38 | 2.4 | |
| **OSA (REI / AHI ≥ 15 )** | | | | | | | | | |
| No | 4167.0 | 82.5 | | 723 | 47.9 | | 1143 | 71.7 | |
| Yes | 882.8 | 17.5 | | 667 | 44.2 | | 287 | 18.0 | |
| Missing | | | | 121 | 8.0 | | 164 | 10.3 | |
| **Education[c]** | | | | | | | | | |
| No High School Diploma | 2307.7 | 40.5 | | 210 | 13.9 | | 172 | 10.8 | |
| At Most a High School Diploma | 1239.9 | 21.7 | | 258 | 17.1 | | 611 | 38.3 | |
| Greater than High School Education | 2153.8 | 37.8 | | 1043 | 69.0 | | 811 | 50.9 | |
| **APOE-4** | | | | | | | | | |
| 0 | 4419.0 | 77.5 | | 1088 | 72.0 | | 1126 | 70.6 | |
| 1 | 1197.9 | 21.0 | | 390 | 25.8 | | 422 | 26.5 | |
| 2 | 84.6 | 1.5 | | 33 | 2.2 | | 46 | 2.9 | |

a) Means, standard deviations (SD), and percentages (%) for HCHS/SOL presented in this table are weighted for the study's sampling weights. For unweighted means, standard Deviations (SD), and percentages (%) see Table 3.
b) Baseline was HCHS/SOL visit 1, MESA visit 5, and ARIC visit 4
c) Follow-up was HCHS/SOL visit 2, MESA visit 6, and ARIC visit 5

**Supplemental Table 2. Study-Specific and Meta-Analysis Results for the Association between Sleep Phenotypes and Global Cognitive Function Measures, M1 Adjustment Set (age, sex, center, education level, race/ethnicity or Hispanic background (for HCHS/SOL), and language of cognitive assessments (for HCHS/SOL and MESA))**

| | | ARIC | | | | | | | HCHS/SOL | | | | | | | MESA 5 | | | | | | | Meta-Analysis (Fixed Effects) | | | | | | | | |
|---|---|---|---|---|---|---|---|---|---|---|---|---|---|---|---|---|---|---|---|---|---|---|---|---|---|---|---|---|---|---|---|
| **Cognitive Phenotype** | **Sleep Phenotype** | **N** | **Beta** | **SE** | **Pvalue** | | | | **N** | **Beta** | **SE** | **Pvalue** | | | | **N** | **Beta** | **SE** | **Pvalue** | | | | **N** | **Beta** | **SE** | **Pvalue** | **Pvalue FDR** | **P for Heterogeneity** | | | |
| GCF_baseline | OSA | 1430 | -0.055 | 0.069 | 0.42 | | | | 7605 | 0.028 | 0.039 | 0.46 | | | | 1390 | -0.053 | 0.037 | 0.15 | | | | 10425 | -0.022 | 0.025 | 0.38 | 0.90 | 0.24 | | | |
| GCF_baseline | Short_Sleep | 1555 | -0.108 | 0.095 | 0.26 | | | | 8137 | 0.042 | 0.044 | 0.34 | | | | 1492 | -0.099 | 0.064 | 0.12 | | | | 11184 | -0.023 | 0.034 | 0.49 | 0.90 | 0.07 | | | |
| GCF_baseline | Long_Sleep | 1555 | -0.173 | 0.113 | 0.12 | | | | 8137 | -0.205 | 0.033 | 1.46E-09 | | | | 1492 | -0.187 | 0.040 | 2.95E-06 | | | | 11184 | -0.190 | 0.025 | 3.78E-14 | 7.56E-13 | 0.78 | | | |
| GCF_baseline | EDS | 1556 | -0.037 | 0.058 | 0.53 | | | | 8363 | -0.059 | 0.039 | 0.13 | | | | 1496 | -0.001 | 0.051 | 0.98 | | | | 11415 | -0.044 | 0.027 | 0.10 | 0.33 | 0.86 | | | |
| GCF_baseline | Insomnia | 1582 | 0.067 | 0.052 | 0.20 | | | | 8272 | -0.111 | 0.029 | 1.35E-04 | | | | 1491 | 0.007 | 0.038 | 0.86 | | | | 11345 | -0.051 | 0.021 | 0.01 | 0.07 | 0.005 | | | |
| GCF_change | OSA | 795 | -0.015 | 0.083 | 0.86 | | | | 5406 | -0.081 | 0.047 | 0.08 | | | | 665 | 0.002 | 0.042 | 0.96 | | | | 6866 | -0.039 | 0.029 | 0.18 | 0.52 | 0.51 | | | |
| GCF_change | Short_Sleep | 879 | -0.006 | 0.115 | 0.96 | | | | 5769 | -0.020 | 0.038 | 0.59 | | | | 708 | 0.062 | 0.069 | 0.36 | | | | 7356 | 0.007 | 0.032 | 0.82 | 0.97 | 0.31 | | | |
| GCF_change | Long_Sleep | 879 | -0.187 | 0.140 | 0.18 | | | | 5769 | -0.013 | 0.040 | 0.75 | | | | 708 | -0.003 | 0.048 | 0.94 | | | | 7356 | -0.023 | 0.030 | 0.45 | 0.90 | 0.48 | | | |
| GCF_change | EDS | 873 | 0.062 | 0.067 | 0.36 | | | | 5899 | 0.031 | 0.039 | 0.43 | | | | 710 | -0.068 | 0.056 | 0.22 | | | | 7482 | 0.009 | 0.028 | 0.76 | 0.97 | 0.23 | | | |
| GCF_change | Insomnia | 886 | -0.052 | 0.061 | 0.40 | | | | 5850 | 0.011 | 0.026 | 0.68 | | | | 709 | 0.003 | 0.042 | 0.95 | | | | 7445 | 0.002 | 0.021 | 0.91 | 0.97 | 0.64 | | | |
| | | **N** | **Beta** | **SE** | **Pvalue** | **OR RR** | **95% CI** | | **N** | **Beta** | **SE** | **Pvalue** | **OR RR** | **95% CI** | | **N** | **Beta** | **SE** | **Pvalue** | **OR RR** | **95% CI** | | **N** | **Beta** | **SE** | **Pvalue** | **Pvalue FDR** | **P for Heterogeneity** | **OR RR** | **95% CI** | |
| Baseline_Poor_GCF | OSA | 1430 | -0.105 | 0.211 | 0.62 | 0.90 | 0.60 | 1.36 | 7605 | -0.016 | 0.123 | 0.90 | 0.98 | 0.77 | 1.25 | 1390 | 0.137 | 0.180 | 0.45 | 1.15 | 0.81 | 1.63 | 10425 | -0.009 | 0.093 | 0.93 | 0.97 | 0.79 | 0.99 | 0.83 | 1.19 |
| Baseline_Poor_GCF | Short_Sleep | 1555 | 0.002 | 0.286 | 0.99 | 1.00 | 0.57 | 1.76 | 8137 | -0.111 | 0.119 | 0.35 | 0.90 | 0.71 | 1.13 | 1492 | 0.501 | 0.255 | 0.05 | 1.65 | 1.00 | 2.72 | 11184 | 0.012 | 0.101 | 0.91 | 0.97 | 0.05 | 1.01 | 0.83 | 1.23 |
| Baseline_Poor_GCF | Long_Sleep | 1555 | -0.020 | 0.351 | 0.95 | 0.98 | 0.49 | 1.95 | 8137 | 0.598 | 0.100 | 3.35E-09 | 1.82 | 1.50 | 2.21 | 1492 | 0.737 | 0.177 | 3.37E-05 | 2.09 | 1.48 | 2.96 | 11184 | 0.570 | 0.086 | 2.96E-11 | 2.96E-10 | 0.22 | 1.77 | 1.49 | 2.09 |
| Baseline_Poor_GCF | EDS | 1556 | -0.154 | 0.182 | 0.40 | 0.86 | 0.60 | 1.23 | 8363 | 0.105 | 0.120 | 0.38 | 1.11 | 0.88 | 1.4 | 1496 | 0.056 | 0.227 | 0.81 | 1.06 | 0.68 | 1.65 | 11415 | 0.038 | 0.092 | 0.68 | 0.97 | 0.48 | 1.04 | 0.87 | 1.24 |
| Baseline_Poor_GCF | Insomnia | 1582 | -0.234 | 0.166 | 0.16 | 0.79 | 0.57 | 1.10 | 8272 | 0.308 | 0.092 | 8.99E-04 | 1.36 | 1.14 | 1.63 | 1491 | 0.138 | 0.174 | 0.43 | 1.15 | 0.82 | 1.62 | 11345 | 0.200 | 0.074 | 6.76E-03 | 0.05 | 0.01 | 1.22 | 1.06 | 1.41 |
| Incident_Poor_GCF | OSA | 732 | 0.301 | 0.268 | 0.26 | 1.35 | 0.80 | 2.29 | 4668 | 0.004 | 0.195 | 0.99 | 0.89 | 0.63 | 1.25 | 596 | 0.064 | 0.228 | 0.78 | 1.07 | 0.68 | 1.67 | 5996 | 0.082 | 0.134 | 0.54 | 0.90 | 0.64 | 1.09 | 0.84 | 1.41 |
| Incident_Poor_GCF | Short_Sleep | 810 | 0.134 | 0.380 | 0.72 | 1.14 | 0.54 | 2.41 | 4964 | -0.074 | 0.188 | 0.69 | 1.01 | 0.68 | 1.5 | 632 | -0.667 | 0.475 | 0.16 | 0.51 | 0.20 | 1.3 | 6406 | -0.104 | 0.161 | 0.52 | 0.90 | 0.33 | 0.90 | 0.66 | 1.24 |
| Incident_Poor_GCF | Long_Sleep | 810 | 0.468 | 0.379 | 0.22 | 1.60 | 0.76 | 3.35 | 4964 | 0.208 | 0.162 | 0.20 | 1.27 | 0.94 | 1.73 | 632 | -0.014 | 0.258 | 0.96 | 0.99 | 0.60 | 1.63 | 6406 | 0.233 | 0.131 | 0.08 | 0.30 | 0.80 | 1.26 | 0.98 | 1.63 |
| Incident_Poor_GCF | EDS | 803 | -0.346 | 0.266 | 0.19 | 0.71 | 0.42 | 1.19 | 5071 | 0.091 | 0.150 | 0.55 | 1.09 | 0.81 | 1.47 | 633 | 0.052 | 0.303 | 0.86 | 1.05 | 0.58 | 1.91 | 6507 | -0.002 | 0.121 | 0.99 | 0.99 | 0.35 | 1.00 | 0.79 | 1.26 |
| Incident_Poor_GCF | Insomnia | 815 | -0.171 | 0.226 | 0.45 | 0.84 | 0.54 | 1.31 | 5027 | -0.003 | 0.119 | 0.98 | 0.98 | 0.76 | 1.26 | 633 | -0.038 | 0.232 | 0.87 | 0.96 | 0.61 | 1.52 | 6475 | -0.042 | 0.097 | 0.67 | 0.97 | 0.81 | 0.96 | 0.79 | 1.16 |

**Supplemental Table 3. Study-Specific and Meta-Analysis Results for the Association between Sleep Phenotypes and Baseline and Change in Global Cognitive Function (GCF) Excluding Individuals with Poor Baseline GCF, M1 Adjustment Set**

| | | ARIC | | | | HCHS/SOL | | | | MESA 5 | | | | Meta-Analysis | | | | |
|---|---|---|---|---|---|---|---|---|---|---|---|---|---|---|---|---|---|---|
| Cognitive Phenotype | Sleep Phenotype | N | Beta | SE | Pvalue | N | Beta | SE | Pvalue | N | Beta | SE | Pvalue | N | Beta | SE | Pvalue | P for Heterogeneity |
| GCF_baseline | OSA | 1264 | -0.115 | 0.061 | 0.06 | 6409 | 0.030 | 0.036 | 0.40 | 1132 | -0.068 | 0.033 | 0.04 | 8805 | -0.036 | 0.022 | 0.11 | 0.05 |
| GCF_baseline | Short_Sleep | 1396 | 0.001 | 0.046 | 0.98 | 6836 | 0.030 | 0.041 | 0.47 | 1217 | -0.005 | 0.059 | 0.93 | 9426 | 0.002 | 0.031 | 0.94 | 0.39 |
| GCF_baseline | Long_Sleep | 1373 | -0.099 | 0.085 | 0.24 | 6836 | -0.108 | 0.029 | 2.41E-04 | 1217 | -0.095 | 0.037 | 0.01 | 9426 | -0.108 | 0.022 | 1.32E-06 | 0.57 |
| GCF_baseline | EDS | 1373 | -0.209 | 0.100 | 0.04 | 7014 | -0.047 | 0.034 | 0.16 | 1219 | -0.043 | 0.045 | 0.34 | 9604 | -0.054 | 0.024 | 0.02 | 0.80 |
| GCF_baseline | Insomnia | 1371 | -0.084 | 0.051 | 0.10 | 6939 | -0.060 | 0.025 | 0.02 | 1217 | 0.025 | 0.033 | 0.45 | 9552 | -0.024 | 0.018 | 0.20 | 0.11 |
| GCF_change | OSA | 732 | 0.015 | 0.086 | 0.86 | 4668 | -0.101 | 0.052 | 0.05 | 596 | -0.019 | 0.041 | 0.64 | 5996 | -0.043 | 0.030 | 0.16 | 0.36 |
| GCF_change | Short_Sleep | 815 | -0.034 | 0.063 | 0.59 | 4964 | 0.008 | 0.044 | 0.85 | 632 | 0.026 | 0.073 | 0.72 | 6406 | 0.010 | 0.036 | 0.78 | 0.95 |
| GCF_change | Long_Sleep | 810 | -0.019 | 0.120 | 0.87 | 4964 | -0.054 | 0.044 | 0.22 | 632 | -0.015 | 0.049 | 0.76 | 6406 | -0.040 | 0.032 | 0.21 | 0.73 |
| GCF_change | EDS | 810 | -0.114 | 0.142 | 0.42 | 5071 | 0.022 | 0.042 | 0.60 | 633 | -0.047 | 0.054 | 0.39 | 6507 | 0.008 | 0.030 | 0.78 | 0.42 |
| GCF_change | Insomnia | 803 | 0.061 | 0.070 | 0.38 | 5027 | 0.003 | 0.029 | 0.92 | 633 | 0.016 | 0.041 | 0.69 | 6475 | 0.002 | 0.022 | 0.92 | 0.80 |

### Supplemental Table 4. Study-Specific and Meta-Analysis Results for the Association between Short and Long Sleep vs Normal Sleep Phenotypes and Global Cognitive Function Measures, M1 Adjustment Set

| | | ARIC | | | | | | | HCHS/SOL | | | | | | | MESA 5 | | | | | | | Meta-Analysis (Fixed Effects) | | | | | | | |
|---|---|---|---|---|---|---|---|---|---|---|---|---|---|---|---|---|---|---|---|---|---|---|---|---|---|---|---|---|---|---|
| **Cognitive Phenotype** | **Sleep Phenotype** | **N** | **Beta** | **SE** | **Pvalue** | | | | **N** | **Beta** | **SE** | **Pvalue** | | | | **N** | **Beta** | **SE** | **Pvalue** | | | | **N** | **Beta** | **SE** | **Pvalue** | **P for Heterogeneity** | | | |
| GCF_baseline | Short_Sleep | 1466 | -0.082 | 0.066 | 0.22 | | | | 6451 | -0.010 | 0.045 | 0.83 | | | | 1136 | -0.160 | 0.061 | 0.009 | | | | 9053 | -0.067 | 0.032 | 0.04 | 0.14 | | | |
| GCF_baseline | Long_Sleep | 1426 | -0.121 | 0.078 | 0.12 | | | | 7249 | -0.206 | 0.034 | 2.00E-09 | | | | 1364 | -0.185 | 0.040 | 4.84E-06 | | | | 10039 | -0.190 | 0.025 | 1.21E-14 | 0.60 | | | |
| GCF_change | Short_Sleep | 836 | -0.008 | 0.083 | 0.92 | | | | 4640 | -0.022 | 0.039 | 0.57 | | | | 571 | 0.084 | 0.069 | 0.22 | | | | 6047 | 0.002 | 0.031 | 0.95 | 0.40 | | | |
| GCF_change | Long_Sleep | 813 | -0.148 | 0.101 | 0.14 | | | | 5145 | -0.016 | 0.041 | 0.70 | | | | 643 | -0.013 | 0.048 | 0.79 | | | | 6601 | -0.026 | 0.030 | 0.38 | 0.45 | | | |
| | | **N** | **Beta** | **SE** | **Pvalue** | **OR RR** | **95% CI** | | **N** | **Beta** | **SE** | **Pvalue** | **OR RR** | **95% CI** | | **N** | **Beta** | **SE** | **Pvalue** | **OR RR** | **95% CI** | | **N** | **Beta** | **SE** | **Pvalue** | **P for Heterogeneity** | **OR RR** | **95% CI** | |
| Baseline_Poor_GCF | Short_Sleep | 1466 | 0.417 | 0.339 | 0.22 | 1.61 | 0.84 | 3.08 | 6451 | 0.066 | 0.120 | 0.58 | 1.07 | 0.84 | 1.35 | 1136 | 0.779 | 0.279 | 0.005 | 2.18 | 1.26 | 3.77 | 9053 | 0.200 | 0.105 | 0.06 | 0.05 | 1.22 | 0.99 | 1.50 |
| Baseline_Poor_GCF | Long_Sleep | 1426 | 0.269 | 0.441 | 0.54 | 1.36 | 0.60 | 3.10 | 7249 | 0.605 | 0.101 | 4.09E-09 | 1.83 | 1.50 | 2.23 | 1364 | 0.762 | 0.196 | 0.0001 | 2.14 | 1.46 | 3.15 | 10039 | 0.624 | 0.088 | 1.62E-12 | 0.55 | 1.87 | 1.57 | 2.22 |
| Incident_Poor_GCF | Short_Sleep | 804 | 0.232 | 0.384 | 0.55 | 1.28 | 0.60 | 2.70 | 4060 | -0.022 | 0.187 | 0.91 | 1.07 | 0.71 | 1.60 | 515 | -0.798 | 0.525 | 0.13 | 0.45 | 0.16 | 1.26 | 5379 | -0.050 | 0.160 | 0.75 | 0.27 | 0.95 | 0.69 | 1.30 |
| Incident_Poor_GCF | Long_Sleep | 784 | 0.646 | 0.365 | 0.08 | 1.83 | 0.89 | 3.75 | 4436 | 0.203 | 0.163 | 0.21 | 1.27 | 0.93 | 1.74 | 586 | 0.146 | 0.282 | 0.60 | 1.16 | 0.67 | 2.01 | 5806 | 0.248 | 0.132 | 0.06 | 0.50 | 1.28 | 0.99 | 1.66 |

**Supplemental Table 5. Study-Specific and Meta-Analysis Results for the Association between Continuous Sleep Phenotypes and Global Cognitive Function Measures, M1 Adjustment Set (age, sex, center, education level, race/ethnicity or Hispanic background (for HCHS/SOL), and language of cognitive assessments (for HCHS/SOL and MESA))**

| | | ARIC | | | | | | | HCHS/SOL | | | | | | | MESA 5 | | | | | | | Meta-Analysis (Fixed Effects) | | | | | | | |
|---|---|---|---|---|---|---|---|---|---|---|---|---|---|---|---|---|---|---|---|---|---|---|---|---|---|---|---|---|---|---|
| **Cognitive Phenotype** | **Sleep Phenotype** | **N** | **Beta** | **SE** | **Pvalue** | | | | **N** | **Beta** | **SE** | **Pvalue** | | | | **N** | **Beta** | **SE** | **Pvalue** | | | | **N** | **Beta** | **SE** | **Pvalue** | **P for Heterogeneity** | | | |
| GCF_baseline | AHI | 1430 | -0.001 | 0.002 | 0.56 | | | | 7605 | 0.001 | 0.001 | 0.25 | | | | 1390 | -0.001 | 0.001 | 0.21 | | | | 10425 | -3.80E-04 | 6.28E-04 | 0.54 | 0.13 | | | |
| GCF_baseline | ESS | 1556 | -0.005 | 0.004 | 0.20 | | | | 8363 | -0.002 | 0.003 | 0.40 | | | | 1496 | -0.002 | 0.004 | 0.70 | | | | 11415 | -0.003 | 0.002 | 0.08 | 0.76 | | | |
| GCF_baseline | WHIIRS | 1582 | 0.005 | 0.007 | 0.51 | | | | 8272 | -0.010 | 0.002 | 6.66E-05 | | | | 1491 | 0.001 | 0.004 | 0.75 | | | | 11345 | -0.005 | 0.002 | 3.49E-03 | 0.02 | | | |
| GCF_baseline | Sleep Duration | 1555 | -0.010 | 0.019 | 0.58 | | | | 8137 | -0.047 | 0.009 | 3.36E-07 | | | | 1492 | -0.021 | 0.013 | 0.10 | | | | 11184 | -0.034 | 0.007 | 6.74E-07 | 0.09 | | | |
| GCF_change | AHI | 795 | 0.000 | 0.002 | 0.91 | | | | 5406 | -0.001 | 0.001 | 0.45 | | | | 665 | 0.000 | 0.001 | 0.95 | | | | 6866 | -3.91E-04 | 7.88E-04 | 0.62 | 0.84 | | | |
| GCF_change | ESS | 873 | 0.007 | 0.005 | 0.20 | | | | 5899 | 0.001 | 0.003 | 0.72 | | | | 710 | -0.005 | 0.005 | 0.36 | | | | 7482 | 8.40E-04 | 2.25E-03 | 0.71 | 0.26 | | | |
| GCF_change | WHIIRS | 886 | -0.016 | 0.009 | 0.08 | | | | 5850 | 8.24E-04 | 0.003 | 0.74 | | | | 709 | 0.004 | 0.004 | 0.31 | | | | 7445 | 2.11E-04 | 2.02E-03 | 0.92 | 0.12 | | | |
| GCF_change | Sleep Duration | 879 | 0.003 | 0.024 | 0.89 | | | | 5769 | -2.96E-04 | 0.010 | 0.98 | | | | 708 | -0.02827 | 0.015 | 0.06 | | | | 7356 | -0.008 | 0.008 | 0.31 | 0.26 | | | |
| | | **N** | **Beta** | **SE** | **Pvalue** | **OR RR** | **95% CI** | | **N** | **Beta** | **SE** | **Pvalue** | **OR RR** | **95% CI** | | **N** | **Beta** | **SE** | **Pvalue** | **OR RR** | **95% CI** | | **N** | **Beta** | **SE** | **Pvalue** | **P for Heterogeneity** | **OR RR** | **95% CI** | |
| Baseline_Poor_GCF | AHI | 1430 | -0.006 | 0.009 | 0.51 | 0.99 | 0.98 | 1.01 | 7605 | -0.001 | 0.003 | 0.81 | 1.00 | 0.99 | 1.01 | 1390 | 0.007 | 0.005 | 0.12 | 1.01 | 1.00 | 1.02 | 10425 | 0.001 | 0.003 | 0.62 | 0.26 | 1.00 | 1.00 | 1.01 |
| Baseline_Poor_GCF | ESS | 1556 | -0.015 | 0.025 | 0.57 | 0.99 | 0.94 | 1.04 | 8363 | -0.001 | 0.009 | 0.91 | 1.00 | 0.98 | 1.02 | 1496 | -0.016 | 0.019 | 0.40 | 0.98 | 0.95 | 1.02 | 11415 | -0.004 | 0.008 | 0.64 | 0.84 | 1.00 | 0.98 | 1.01 |
| Baseline_Poor_GCF | WHIIRS | 1582 | -0.013 | 0.045 | 0.78 | 0.99 | 0.90 | 1.08 | 8272 | 0.025 | 0.008 | 1.82E-03 | 1.03 | 1.01 | 1.04 | 1491 | 0.015 | 0.018 | 0.41 | 1.01 | 0.98 | 1.05 | 11345 | 0.024 | 0.007 | 8.26E-04 | 0.47 | 1.02 | 1.01 | 1.04 |
| Baseline_Poor_GCF | Sleep Duration | 1555 | 0.079 | 0.110 | 0.48 | 1.08 | 0.87 | 1.34 | 8137 | 0.136 | 0.030 | 8.52E-06 | 1.15 | 1.08 | 1.22 | 1492 | 0.046 | 0.058 | 0.42 | | | | 11184 | 0.115 | 0.026 | 1.12E-05 | 0.37 | 1.12 | 1.07 | 1.18 |
| Incident_Poor_GCF | AHI | 766 | -0.004 | 0.009 | 0.68 | 1.00 | 0.98 | 1.01 | 4668 | -0.006 | 0.006 | 0.25 | 0.99 | 0.98 | 1.00 | 596 | 0.004 | 0.006 | 0.45 | 1.00 | 0.99 | 1.02 | 6030 | -0.003 | 0.004 | 0.43 | 0.62 | 1.00 | 0.99 | 1.00 |
| Incident_Poor_GCF | ESS | 841 | -0.030 | 0.025 | 0.24 | 0.97 | 0.92 | 1.02 | 5071 | 0.007 | 0.013 | 0.59 | 1.01 | 0.98 | 1.03 | 633 | -0.031 | 0.029 | 0.28 | 0.97 | 0.92 | 1.03 | 6545 | -0.003 | 0.011 | 0.78 | 0.35 | 1.00 | 0.98 | 1.02 |
| Incident_Poor_GCF | WHIIRS | 853 | 0.001 | 0.045 | 0.98 | 1.00 | 0.92 | 1.09 | 5027 | 0.009 | 0.011 | 0.41 | 1.01 | 0.99 | 1.04 | 633 | -0.007 | 0.023 | 0.77 | 0.99 | 0.95 | 1.04 | 6513 | 0.005 | 0.010 | 0.62 | 0.69 | 1.00 | 0.99 | 1.02 |
| Incident_Poor_GCF | Sleep Duration | 847 | -0.062 | 0.113 | 0.58 | 0.94 | 0.75 | 1.17 | 4964 | 0.040 | 0.050 | 0.42 | 1.03 | 0.93 | 1.14 | 632 | 0.106 | 0.080 | 0.18 | | | | 6443 | 0.044 | 0.039 | 0.27 | 0.47 | 1.04 | 0.97 | 1.13 |

**Supplemental Table 6. Study-Specific and Meta-Analysis Results for the Association between Sleep Phenotypes and Global Cognitive Function Measures, M2 Adjustment Set (all variables from M1 plus hypertension (defined as >=140/90 or med use), diabetes, body mass index (BMI), high-density lipoprotein (HDL), low-density lipoprotein (LDL) and estimate glomerular filtration rate (eGFR))**

| | | ARIC | | | | | | | HCHS/SOL | | | | | | | MESA 5 | | | | | | | Meta-Analysis (Fixed Effects) | | | | | | | |
|---|---|---|---|---|---|---|---|---|---|---|---|---|---|---|---|---|---|---|---|---|---|---|---|---|---|---|---|---|---|---|
| Cognitive Phenotype | Sleep Phenotype | N | Beta | SE | Pvalue | | | | N | Beta | SE | Pvalue | | | | N | Beta | SE | Pvalue | | | | N | Beta | SE | Pvalue | P for Heterogeneity | | | |
| GCF_baseline | OSA | 1403 | -0.004 | 0.074 | 0.96 | | | | 7344 | 0.062 | 0.042 | 0.15 | | | | 1367 | -0.029 | 0.039 | 0.45 | | | | 10114 | 0.011 | 0.027 | 0.68 | 0.30 | | | |
| GCF_baseline | Short_Sleep | 1524 | -0.083 | 0.096 | 0.38 | | | | 7872 | 0.035 | 0.044 | 0.43 | | | | 1467 | -0.082 | 0.064 | 0.20 | | | | 10863 | -0.025 | 0.034 | 0.47 | 0.11 | | | |
| GCF_baseline | Long_Sleep | 1524 | -0.157 | 0.114 | 0.17 | | | | 7872 | -0.195 | 0.033 | 9.12E-09 | | | | 1467 | -0.179 | 0.040 | 6.99E-06 | | | | 10863 | -0.179 | 0.025 | 1.05E-12 | 0.76 | | | |
| GCF_baseline | EDS | 1527 | -0.021 | 0.059 | 0.72 | | | | 8082 | -0.053 | 0.037 | 0.16 | | | | 1471 | 0.012 | 0.050 | 0.81 | | | | 11080 | -0.034 | 0.027 | 0.20 | 0.77 | | | |
| GCF_baseline | Insomnia | 1552 | 0.058 | 0.053 | 0.27 | | | | 7991 | -0.097 | 0.029 | 9.72E-04 | | | | 1466 | -0.005 | 0.037 | 0.88 | | | | 11009 | -0.047 | 0.021 | 0.02 | 0.02 | | | |
| GCF_change | OSA | 781 | 0.018 | 0.090 | 0.84 | | | | 5218 | -0.106 | 0.046 | 0.02 | | | | 655 | -0.010 | 0.045 | 0.83 | | | | 6654 | -0.055 | 0.030 | 0.07 | 0.31 | | | |
| GCF_change | Short_Sleep | 865 | -0.011 | 0.115 | 0.92 | | | | 5574 | -0.020 | 0.038 | 0.60 | | | | 698 | 0.060 | 0.069 | 0.39 | | | | 7137 | 0.006 | 0.032 | 0.84 | 0.33 | | | |
| GCF_change | Long_Sleep | 865 | -0.177 | 0.142 | 0.21 | | | | 5574 | -7.00E-04 | 0.041 | 0.99 | | | | 698 | -0.007 | 0.049 | 0.89 | | | | 7137 | -0.017 | 0.031 | 0.58 | 0.48 | | | |
| GCF_change | EDS | 860 | 0.064 | 0.068 | 0.35 | | | | 5698 | 0.029 | 0.039 | 0.46 | | | | 700 | -0.067 | 0.056 | 0.23 | | | | 7258 | 0.008 | 0.029 | 0.78 | 0.24 | | | |
| GCF_change | Insomnia | 872 | -0.054 | 0.062 | 0.38 | | | | 5649 | 0.008 | 0.027 | 0.76 | | | | 699 | -0.005 | 0.043 | 0.91 | | | | 7220 | -0.001 | 0.021 | 0.98 | 0.65 | | | |
| | | N | Beta | SE | Pvalue | OR RR | 95% CI | | N | Beta | SE | Pvalue | OR RR | 95% CI | | N | Beta | SE | Pvalue | OR RR | 95% CI | | N | Beta | SE | Pvalue | P for Heterogeneity | OR | 95% CI | |
| Baseline_Poor_GCF | OSA | 1403 | -0.018 | 0.226 | 0.94 | 0.98 | 0.63 | 1.53 | 7344 | -0.045 | 0.133 | 0.73 | 0.96 | 0.74 | 1.24 | 1367 | 0.066 | 0.193 | 0.73 | 1.07 | 0.73 | 1.56 | 10114 | -0.029 | 0.100 | 0.77 | 0.98 | 0.97 | 0.80 | 1.18 |
| Baseline_Poor_GCF | Short_Sleep | 1524 | -0.017 | 0.291 | 0.95 | 0.98 | 0.56 | 1.74 | 7872 | -0.088 | 0.122 | 0.47 | 0.92 | 0.72 | 1.16 | 1467 | 0.444 | 0.261 | 0.09 | 1.56 | 0.93 | 2.6 | 10863 | 0.027 | 0.104 | 0.79 | 0.06 | 1.03 | 0.84 | 1.26 |
| Baseline_Poor_GCF | Long_Sleep | 1524 | 0.030 | 0.357 | 0.93 | 1.03 | 0.51 | 2.08 | 7872 | 0.563 | 0.098 | 1.40E-08 | 1.76 | 1.45 | 2.13 | 1467 | 0.738 | 0.182 | 0.00 | 2.09 | 1.46 | 2.99 | 10863 | 0.542 | 0.085 | 1.83E-10 | 0.33 | 1.72 | 1.46 | 2.03 |
| Baseline_Poor_GCF | EDS | 1527 | -0.165 | 0.185 | 0.37 | 0.85 | 0.59 | 1.22 | 8082 | 0.115 | 0.117 | 0.33 | 1.12 | 0.89 | 1.41 | 1471 | -0.028 | 0.232 | 0.90 | 0.97 | 0.62 | 1.53 | 11080 | 0.032 | 0.092 | 0.73 | 0.44 | 1.03 | 0.86 | 1.24 |
| Baseline_Poor_GCF | Insomnia | 1552 | -0.205 | 0.168 | 0.22 | 0.81 | 0.59 | 1.13 | 7991 | 0.278 | 0.097 | 4.23E-03 | 1.32 | 1.09 | 1.6 | 1466 | 0.201 | 0.178 | 0.26 | 1.22 | 0.86 | 1.73 | 11009 | 0.189 | 0.077 | 0.01 | 0.03 | 1.21 | 1.04 | 1.40 |
| Incident_Poor_GCF | OSA | 719 | 0.142 | 0.305 | 0.64 | 1.15 | 0.63 | 2.10 | 4509 | -0.094 | 0.188 | 0.62 | 0.82 | 0.58 | 1.17 | 587 | 0.025 | 0.216 | 0.91 | 1.03 | 0.67 | 1.57 | 5815 | -0.019 | 0.135 | 0.89 | 0.80 | 0.98 | 0.75 | 1.28 |
| Incident_Poor_GCF | Short_Sleep | 797 | 0.199 | 0.386 | 0.61 | 1.22 | 0.57 | 2.60 | 4800 | -0.169 | 0.197 | 0.39 | 0.90 | 0.6 | 1.36 | 623 | -0.672 | 0.453 | 0.14 | 0.51 | 0.21 | 1.24 | 6220 | -0.171 | 0.166 | 0.30 | 0.26 | 0.84 | 0.61 | 1.17 |
| Incident_Poor_GCF | Long_Sleep | 797 | 0.394 | 0.393 | 0.32 | 1.48 | 0.69 | 3.21 | 4800 | 0.204 | 0.165 | 0.22 | 1.26 | 0.93 | 1.7 | 623 | -0.129 | 0.260 | 0.62 | 0.88 | 0.53 | 1.46 | 6220 | 0.199 | 0.134 | 0.14 | 0.81 | 1.22 | 0.94 | 1.59 |
| Incident_Poor_GCF | EDS | 791 | -0.471 | 0.285 | 0.10 | 0.62 | 0.36 | 1.09 | 4900 | 0.070 | 0.156 | 0.65 | 1.09 | 0.81 | 1.48 | 624 | 0.049 | 0.293 | 0.87 | 1.05 | 0.59 | 1.86 | 6315 | -0.033 | 0.125 | 0.79 | 0.23 | 0.97 | 0.76 | 1.24 |
| Incident_Poor_GCF | Insomnia | 802 | -0.156 | 0.235 | 0.51 | 0.86 | 0.54 | 1.36 | 4858 | 0.010 | 0.123 | 0.93 | 1.01 | 0.78 | 1.30 | 624 | 0.007 | 0.224 | 0.97 | 1.01 | 0.65 | 1.56 | 6284 | -0.024 | 0.099 | 0.81 | 0.82 | 0.98 | 0.80 | 1.19 |

**Supplemental Table 7. Study-Specific and Meta-Analysis Results for the Association between Sleep Phenotypes and Global Cognitive Function Measures, M3 Adjustment Set (all variables from M2 plus cigarette use and alcohol use)**

| | | ARIC | | | | | | | HCHS/SOL | | | | | | | MESA 5 | | | | | | | Meta-Analysis (Fixed Effects) | | | | | | | |
|---|---|---|---|---|---|---|---|---|---|---|---|---|---|---|---|---|---|---|---|---|---|---|---|---|---|---|---|---|---|---|
| Cognitive Phenotype | Sleep Phenotype | N | Beta | SE | Pvalue | | | | N | Beta | SE | Pvalue | | | | N | Beta | SE | Pvalue | | | | N | Beta | SE | Pvalue | P for Heterogeneity | | | |
| GCF_baseline | OSA | 1403 | 0.013 | 0.074 | 0.86 | | | | 7329 | 0.058 | 0.041 | 0.16 | | | | 1361 | -0.044 | 0.038 | 0.25 | | | | 10093 | 0.005 | 0.026 | 0.83 | 0.21 | | | |
| GCF_baseline | Short_Sleep | 1524 | -0.075 | 0.095 | 0.43 | | | | 7860 | 0.048 | 0.043 | 0.26 | | | | 1460 | -0.085 | 0.063 | 0.18 | | | | 10844 | -0.013 | 0.033 | 0.70 | 0.09 | | | |
| GCF_baseline | Long_Sleep | 1524 | -0.138 | 0.114 | 0.22 | | | | 7860 | -0.194 | 0.033 | 4.79E-09 | | | | 1460 | -0.162 | 0.040 | 4.54E-05 | | | | 10844 | -0.172 | 0.025 | 2.95E-12 | 0.60 | | | |
| GCF_baseline | EDS | 1527 | -0.020 | 0.059 | 0.74 | | | | 8070 | -0.048 | 0.037 | 0.19 | | | | 1464 | 0.012 | 0.050 | 0.81 | | | | 11061 | -0.031 | 0.026 | 0.24 | 0.80 | | | |
| GCF_baseline | Insomnia | 1552 | 0.062 | 0.052 | 0.24 | | | | 7979 | -0.093 | 0.029 | 1.14E-03 | | | | 1459 | -0.021 | 0.037 | 0.57 | | | | 10990 | -0.049 | 0.021 | 0.02 | 0.03 | | | |
| GCF_change | OSA | 781 | 0.028 | 0.090 | 0.76 | | | | 5207 | -0.109 | 0.045 | 0.02 | | | | 652 | -0.012 | 0.045 | 0.80 | | | | 6640 | -0.057 | 0.030 | 0.06 | 0.26 | | | |
| GCF_change | Short_Sleep | 865 | 0.003 | 0.115 | 0.98 | | | | 5565 | -0.021 | 0.038 | 0.57 | | | | 695 | 0.061 | 0.071 | 0.39 | | | | 7125 | 0.005 | 0.032 | 0.86 | 0.33 | | | |
| GCF_change | Long_Sleep | 865 | -0.172 | 0.141 | 0.22 | | | | 5565 | -0.003 | 0.041 | 0.94 | | | | 695 | -0.005 | 0.050 | 0.93 | | | | 7125 | -0.017 | 0.031 | 0.59 | 0.52 | | | |
| GCF_change | EDS | 860 | 0.060 | 0.068 | 0.38 | | | | 5689 | 0.024 | 0.039 | 0.54 | | | | 697 | -0.066 | 0.056 | 0.24 | | | | 7246 | 0.006 | 0.029 | 0.85 | 0.28 | | | |
| GCF_change | Insomnia | 872 | -0.053 | 0.061 | 0.39 | | | | 5640 | 0.012 | 0.026 | 0.65 | | | | 696 | -0.002 | 0.043 | 0.96 | | | | 7208 | 0.002 | 0.021 | 0.91 | 0.63 | | | |
| | | N | Beta | SE | Pvalue | OR RR | 95% CI | | N | Beta | SE | Pvalue | OR RR | 95% CI | | N | Beta | SE | Pvalue | OR RR | 95% CI | | N | Beta | SE | Pvalue | P for Heterogeneity | OR | 95% CI | |
| Baseline_Poor_GCF | OSA | 1403 | -0.044 | 0.225 | 0.84 | 0.96 | 0.62 | 1.49 | 7329 | -0.026 | 0.133 | 0.84 | 0.97 | 0.75 | 1.26 | 1361 | 0.157 | 0.196 | 0.42 | 1.17 | 0.8 | 1.72 | 10093 | 0.001 | 0.100 | 1.00 | 0.85 | 1.00 | 0.82 | 1.22 |
| Baseline_Poor_GCF | Short_Sleep | 1524 | -0.017 | 0.290 | 0.95 | 0.98 | 0.56 | 1.74 | 7860 | -0.114 | 0.124 | 0.36 | 0.89 | 0.7 | 1.14 | 1460 | 0.462 | 0.266 | 0.08 | 1.59 | 0.94 | 2.67 | 10844 | 0.008 | 0.105 | 0.94 | 0.06 | 1.01 | 0.82 | 1.24 |
| Baseline_Poor_GCF | Long_Sleep | 1524 | 0.013 | 0.359 | 0.97 | 1.01 | 0.50 | 2.05 | 7860 | 0.573 | 0.098 | 9.72E-09 | 1.77 | 1.46 | 2.15 | 1460 | 0.728 | 0.185 | 8.28E-05 | 2.07 | 1.44 | 2.97 | 10844 | 0.548 | 0.085 | 1.40E-10 | 0.30 | 1.73 | 1.46 | 2.05 |
| Baseline_Poor_GCF | EDS | 1527 | -0.170 | 0.184 | 0.36 | 0.84 | 0.59 | 1.21 | 8070 | 0.109 | 0.116 | 0.35 | 1.12 | 0.89 | 1.4 | 1464 | -0.033 | 0.234 | 0.89 | 0.97 | 0.61 | 1.53 | 11061 | 0.025 | 0.091 | 0.78 | 0.44 | 1.03 | 0.86 | 1.23 |
| Baseline_Poor_GCF | Insomnia | 1552 | -0.218 | 0.168 | 0.20 | 0.80 | 0.58 | 1.12 | 7979 | 0.269 | 0.098 | 6.25E-03 | 1.31 | 1.08 | 1.59 | 1459 | 0.254 | 0.181 | 0.16 | 1.29 | 0.91 | 1.84 | 10990 | 0.186 | 0.077 | 0.02 | 0.02 | 1.20 | 1.04 | 1.40 |
| Incident_Poor_GCF | OSA | 719 | 0.188 | 0.306 | 0.54 | 1.21 | 0.66 | 2.20 | 4500 | -0.104 | 0.180 | 0.56 | 0.82 | 0.58 | 1.15 | 584 | -0.005 | 0.216 | 0.98 | 0.99 | 0.65 | 1.52 | 5803 | -0.018 | 0.131 | 0.89 | 0.71 | 0.98 | 0.76 | 1.27 |
| Incident_Poor_GCF | Short_Sleep | 797 | 0.198 | 0.389 | 0.61 | 1.22 | 0.57 | 2.61 | 4792 | -0.186 | 0.191 | 0.33 | 0.88 | 0.59 | 1.32 | 620 | -0.615 | 0.454 | 0.18 | 0.54 | 0.22 | 1.32 | 6209 | -0.185 | 0.163 | 0.26 | 0.27 | 0.83 | 0.60 | 1.14 |
| Incident_Poor_GCF | Long_Sleep | 797 | 0.409 | 0.394 | 0.30 | 1.51 | 0.70 | 3.26 | 4792 | 0.219 | 0.159 | 0.17 | 1.29 | 0.96 | 1.72 | 620 | -0.131 | 0.259 | 0.61 | 0.88 | 0.53 | 1.46 | 6209 | 0.211 | 0.130 | 0.10 | 0.80 | 1.24 | 0.96 | 1.59 |
| Incident_Poor_GCF | EDS | 791 | -0.454 | 0.286 | 0.11 | 0.64 | 0.36 | 1.11 | 4892 | 0.084 | 0.157 | 0.59 | 1.11 | 0.82 | 1.5 | 621 | -0.032 | 0.293 | 0.91 | 0.97 | 0.55 | 1.72 | 6304 | -0.034 | 0.125 | 0.79 | 0.25 | 0.97 | 0.76 | 1.24 |
| Incident_Poor_GCF | Insomnia | 802 | -0.165 | 0.236 | 0.48 | 0.85 | 0.53 | 1.35 | 4850 | 0.004 | 0.117 | 0.97 | 1.00 | 0.79 | 1.26 | 621 | 0.026 | 0.223 | 0.91 | 1.03 | 0.66 | 1.59 | 6273 | -0.026 | 0.096 | 0.79 | 0.81 | 0.97 | 0.81 | 1.18 |

**Supplemental Table 8. Meta-Analysis Results for the Association between Sleep Phenotypes and Individual Cognitive Tests, M1 Adjustment Set (age, sex, center, education level, race/ethnicity or Hispanic background (for HCHS/SOL), and language of cognitive assessments (for HCHS/SOL and MESA))**

| Cognitive Phenotype | Sleep Phenotype | N | Beta | SE | Pvalue | Pval FDR | Pval Heterogeneity |
|---|---|---|---|---|---|---|---|
| DSS_baseline | Long_Sleep | 11315 | -0.166 | 0.022 | 4.83E-14 | 2.90E-12 | 0.37 |
| CASI/SIS_baseline | Long_Sleep | 9991 | -0.163 | 0.026 | 8.17E-10 | 2.45E-08 | 0.75 |
| SEVLT_3Trial_baseline | Insomnia | 8600 | -0.130 | 0.028 | 3.12E-06 | 6.25E-05 | 1.00 |
| SEVLT/DWRT_baseline | Long_Sleep | 10028 | -0.137 | 0.031 | 9.17E-06 | 1.38E-04 | 0.44 |
| SEVLT_3Trial_baseline | Long_Sleep | 8463 | -0.130 | 0.034 | 1.29E-04 | 1.55E-03 | 1.00 |
| WF_baseline | Long_Sleep | 9899 | -0.128 | 0.038 | 8.79E-04 | 0.009 | 0.83 |
| SEVLT/DWRT_baseline | Insomnia | 10191 | -0.066 | 0.023 | 0.004 | 0.03 | 0.24 |
| DSS_baseline | Insomnia | 11478 | -0.048 | 0.017 | 0.006 | 0.04 | 0.002 |
| DigitSpan_change | EDS | 768 | -0.171 | 0.075 | 0.02 | 0.15 | 1.00 |
| DSS_baseline | EDS | 11549 | -0.051 | 0.023 | 0.02 | 0.15 | 0.35 |
| DSS_change | OSA | 6983 | -0.047 | 0.022 | 0.03 | 0.16 | 0.20 |
| SEVLT_3Trial_baseline | EDS | 8695 | -0.072 | 0.034 | 0.03 | 0.16 | 1.00 |
| DigitSpan_baseline | Long_Sleep | 1492 | -0.103 | 0.049 | 0.04 | 0.16 | 1.00 |
| SEVLT_3Trial_change | Long_Sleep | 6023 | -0.084 | 0.045 | 0.06 | 0.25 | 1.00 |
| WF_baseline | OSA | 9216 | -0.067 | 0.036 | 0.06 | 0.25 | 0.81 |
| SEVLT_3Trial_change | OSA | 5643 | -0.091 | 0.052 | 0.08 | 0.28 | 1.00 |
| SEVLT_3Trial_change | Insomnia | 6105 | 0.057 | 0.033 | 0.08 | 0.28 | 1.00 |
| DigitSpan_change | OSA | 720 | -0.097 | 0.057 | 0.09 | 0.28 | 1.00 |
| SEVLT/DWRT_change | Short_Sleep | 6910 | -0.093 | 0.056 | 0.09 | 0.28 | 0.19 |
| DSS_baseline | OSA | 10544 | -0.037 | 0.022 | 0.10 | 0.28 | 0.04 |
| SEVLT/DWRT_baseline | Short_Sleep | 10028 | 0.061 | 0.037 | 0.10 | 0.28 | 0.60 |
| CASI/SIS_change | OSA | 6407 | -0.083 | 0.051 | 0.10 | 0.28 | 0.69 |
| WF_change | Long_Sleep | 6834 | -0.051 | 0.033 | 0.13 | 0.33 | 0.45 |
| DSS_change | Long_Sleep | 7479 | 0.033 | 0.023 | 0.15 | 0.38 | 0.48 |
| CASI/SIS_baseline | Short_Sleep | 9991 | -0.056 | 0.039 | 0.16 | 0.38 | 0.66 |
| WF_change | EDS | 6961 | 0.037 | 0.028 | 0.19 | 0.44 | 5.64E-03 |
| WF_change | Insomnia | 6921 | -0.029 | 0.024 | 0.23 | 0.49 | 0.75 |
| SEVLT/DWRT_change | OSA | 6445 | -0.054 | 0.046 | 0.23 | 0.49 | 0.67 |
| DigitSpan_baseline | Short_Sleep | 1492 | -0.088 | 0.075 | 0.24 | 0.49 | 1.00 |
| CASI/SIS_baseline | OSA | 9318 | 0.030 | 0.026 | 0.25 | 0.49 | 0.07 |
| DigitSpan_baseline | EDS | 1496 | 0.062 | 0.059 | 0.30 | 0.57 | 1.00 |
| CASI/SIS_change | Short_Sleep | 6836 | 0.062 | 0.069 | 0.37 | 0.66 | 0.19 |
| SEVLT_3Trial_change | Short_Sleep | 6023 | 0.044 | 0.049 | 0.37 | 0.66 | 1.00 |
| WF_baseline | Short_Sleep | 9899 | -0.039 | 0.044 | 0.38 | 0.66 | 0.68 |
| SEVLT_3Trial_baseline | OSA | 7895 | 0.036 | 0.043 | 0.41 | 0.66 | 1.00 |
| SEVLT/DWRT_baseline | OSA | 9333 | 0.026 | 0.033 | 0.42 | 0.66 | 0.40 |
| SEVLT_3Trial_baseline | Short_Sleep | 8463 | 0.033 | 0.043 | 0.44 | 0.66 | 1.00 |
| SEVLT_3Trial_change | EDS | 6157 | 0.035 | 0.046 | 0.44 | 0.66 | 1.00 |
| DSS_change | Short_Sleep | 7479 | -0.018 | 0.023 | 0.44 | 0.66 | 0.75 |
| CASI/SIS_change | Long_Sleep | 6836 | 0.037 | 0.049 | 0.45 | 0.66 | 0.47 |
| CASI/SIS_change | Insomnia | 6919 | -0.029 | 0.039 | 0.45 | 0.66 | 0.24 |
| CASI/SIS_baseline | Insomnia | 10131 | -0.016 | 0.022 | 0.46 | 0.66 | 8.73E-03 |
| SEVLT/DWRT_change | Long_Sleep | 6910 | -0.029 | 0.046 | 0.53 | 0.74 | 0.06 |
| WF_change | OSA | 6373 | 0.021 | 0.035 | 0.55 | 0.74 | 0.67 |

| | | | | | | | |
|---|---|---|---|---|---|---|---|
| DigitSpan_change | Short_Sleep | 767 | 0.053 | 0.091 | 0.56 | 0.74 | 1.00 |
| CASI/SIS_baseline | EDS | 10231 | -0.015 | 0.030 | 0.61 | 0.79 | 0.34 |
| DSS_change | EDS | 7606 | 0.010 | 0.020 | 0.63 | 0.80 | 0.94 |
| WF_baseline | EDS | 10131 | -0.016 | 0.034 | 0.64 | 0.80 | 0.39 |
| DigitSpan_change | Long_Sleep | 767 | 0.029 | 0.066 | 0.66 | 0.81 | 1.00 |
| SEVLT/DWRT_change | EDS | 7038 | -0.014 | 0.044 | 0.75 | 0.89 | 0.33 |
| DSS_baseline | Short_Sleep | 11315 | -0.008 | 0.028 | 0.77 | 0.89 | 0.03 |
| WF_baseline | Insomnia | 10061 | 0.008 | 0.028 | 0.78 | 0.89 | 0.12 |
| DigitSpan_baseline | OSA | 1390 | 0.012 | 0.044 | 0.79 | 0.89 | 1.00 |
| CASI/SIS_change | EDS | 6973 | 0.011 | 0.056 | 0.84 | 0.94 | 0.03 |
| SEVLT/DWRT_change | Insomnia | 6998 | 0.003 | 0.033 | 0.92 | 0.98 | 0.88 |
| DigitSpan_baseline | Insomnia | 1491 | 0.004 | 0.044 | 0.94 | 0.98 | 1.00 |
| DigitSpan_change | Insomnia | 768 | -0.004 | 0.057 | 0.94 | 0.98 | 1.00 |
| WF_change | Short_Sleep | 6834 | -0.003 | 0.041 | 0.94 | 0.98 | 0.36 |
| DSS_change | Insomnia | 7568 | -0.001 | 0.016 | 0.97 | 0.98 | 0.08 |
| SEVLT/DWRT_baseline | EDS | 10261 | -6.88E-05 | 0.029 | 1.00 | 1.00 | 0.38 |

**Supplemental Table 9. Study-Specific and Meta-Analysis Results for the Association between Sleep Phenotypes and Global Cognitive Function Change and Incident Poor Global Cognitive Function, using Inverse Probability Weighting (IPW) and M1 Adjustment Set**

| | | ARIC | | | | | | | HCHS/SOL | | | | | | | MESA 5 | | | | | | | Meta-Analysis (Fixed Effects) | | | | | | | |
|---|---|---|---|---|---|---|---|---|---|---|---|---|---|---|---|---|---|---|---|---|---|---|---|---|---|---|---|---|---|---|
| **Cognitive Phenotype** | **Sleep Phenotype** | **N** | **Beta** | **SE** | **Pvalue** | | | | **N** | **Beta** | **SE** | **Pvalue** | | | | **N** | **Beta** | **SE** | **Pvalue** | | | | **N** | **Beta** | **SE** | **Pvalue** | **P for Heterogeneity** | | | |
| GCF_change | OSA | 795 | -0.0003 | 0.060 | 1.00 | | | | 5406 | -0.042 | 0.041 | 0.30 | | | | 665 | 0.002 | 0.042 | 0.96 | | | | 6866 | -0.027 | 0.028 | 0.32 | 0.36 | | | |
| GCF_change | Short_Sleep | 879 | -0.004 | 0.082 | 0.96 | | | | 5769 | -0.024 | 0.037 | 0.51 | | | | 708 | 0.062 | 0.069 | 0.36 | | | | 7356 | -0.001 | 0.031 | 0.96 | 0.57 | | | |
| GCF_change | Long_Sleep | 879 | -0.144 | 0.101 | 0.15 | | | | 5769 | -0.011 | 0.037 | 0.75 | | | | 708 | -0.003 | 0.048 | 0.94 | | | | 7356 | -0.021 | 0.029 | 0.49 | 0.44 | | | |
| GCF_change | EDS | 873 | 0.041 | 0.048 | 0.40 | | | | 5899 | -0.009 | 0.040 | 0.81 | | | | 710 | -0.068 | 0.056 | 0.22 | | | | 7482 | 0.011 | 0.027 | 0.67 | 0.26 | | | |
| GCF_change | Insomnia | 886 | -0.043 | 0.044 | 0.32 | | | | 5850 | -0.011 | 0.025 | 0.65 | | | | 709 | 0.003 | 0.042 | 0.95 | | | | 7445 | -0.002 | 0.020 | 0.92 | 0.57 | | | |
| | | **N** | **Beta** | **SE** | **Pvalue** | **OR RR** | **95% CI** | | **N** | **Beta** | **SE** | **Pvalue** | **OR RR** | **95% CI** | | **N** | **Beta** | **SE** | **Pvalue** | **OR RR** | **95% CI** | | **N** | **Beta** | **SE** | **Pvalue** | **P for Heterogeneity** | **OR RR** | **95% CI** | |
| Incident_Poor_GCF | OSA | 766 | 0.201 | 0.273 | 0.46 | 1.22 | 0.72 | 2.09 | 4668 | -0.117 | 0.172 | 0.49 | 0.89 | 0.63 | 1.25 | 596 | 0.064 | 0.228 | 0.78 | 1.07 | 0.68 | 1.67 | 6030 | 0.069 | 0.130 | 0.60 | 0.84 | 1.07 | 0.83 | 1.38 |
| Incident_Poor_GCF | Short_Sleep | 847 | 0.153 | 0.380 | 0.69 | 1.17 | 0.55 | 2.45 | 4964 | 0.008 | 0.203 | 0.97 | 1.01 | 0.68 | 1.5 | 632 | -0.667 | 0.475 | 0.16 | 0.51 | 0.20 | 1.30 | 6443 | -0.101 | 0.159 | 0.53 | 0.39 | 0.90 | 0.66 | 1.23 |
| Incident_Poor_GCF | Long_Sleep | 847 | 0.637 | 0.358 | 0.08 | 1.89 | 0.94 | 3.82 | 4964 | 0.240 | 0.156 | 0.12 | 1.27 | 0.94 | 1.73 | 632 | -0.014 | 0.258 | 0.96 | 0.99 | 0.59 | 1.64 | 6443 | 0.208 | 0.128 | 0.11 | 0.34 | 1.23 | 0.96 | 1.58 |
| Incident_Poor_GCF | EDS | 841 | -0.306 | 0.260 | 0.24 | 0.74 | 0.44 | 1.23 | 5071 | 0.087 | 0.153 | 0.57 | 1.09 | 0.81 | 1.47 | 633 | 0.052 | 0.303 | 0.86 | 1.05 | 0.58 | 1.91 | 6545 | 0.001 | 0.120 | 0.99 | 0.41 | 1.00 | 0.79 | 1.27 |
| Incident_Poor_GCF | Insomnia | 853 | -0.440 | 0.231 | 0.06 | 0.64 | 0.41 | 1.01 | 5027 | -0.022 | 0.127 | 0.86 | 0.98 | 0.76 | 1.26 | 633 | -0.038 | 0.232 | 0.87 | 0.96 | 0.61 | 1.52 | 6513 | -0.084 | 0.096 | 0.38 | 0.24 | 0.92 | 0.76 | 1.11 |

**Supplemental Figure 1. Principal Component (PC) Loadings for Global Cognitive Function (GCF)**

**Panel A. HCHS/SOL**

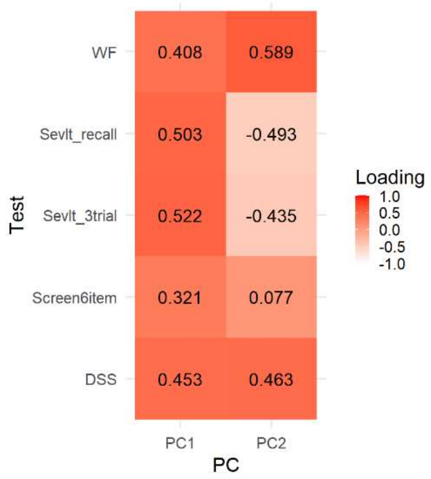


**Panel B. MESA**

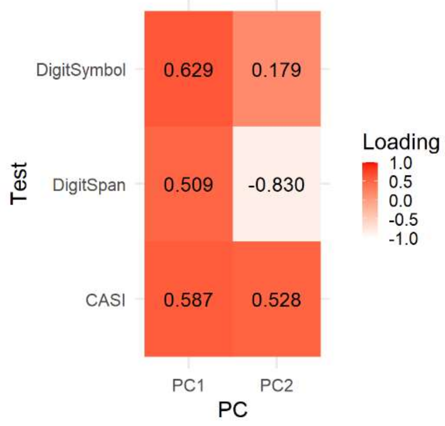


**Panel C. ARIC**

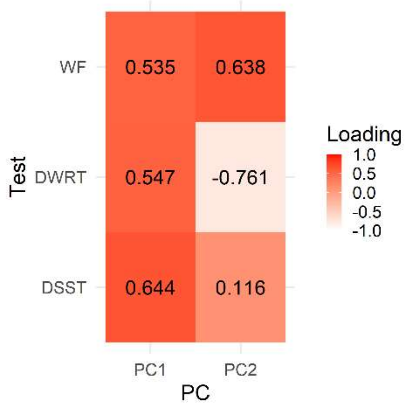

**Supplemental Figure 2. Percent Variance Explained by PCs**

**Panel A. HCHS/SOL**

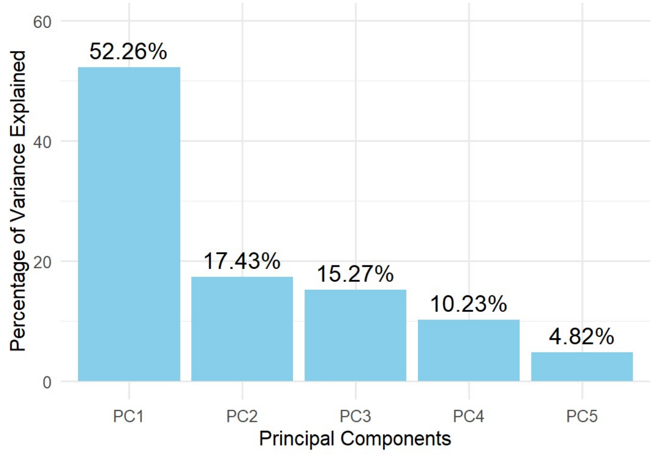


**Panel B. MESA**

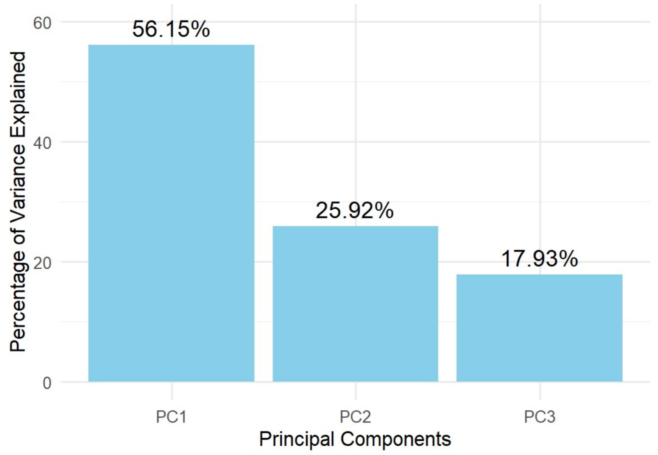


**Panel C. ARIC**

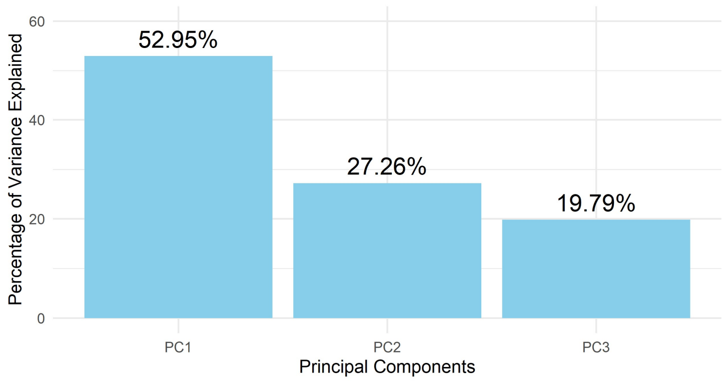

**Supplemental Figure 3. Correlation between GCF (PC1) and Individual Cognitive Assessments**

**Panel A. HCHS/SOL Baseline (Visit 1)**

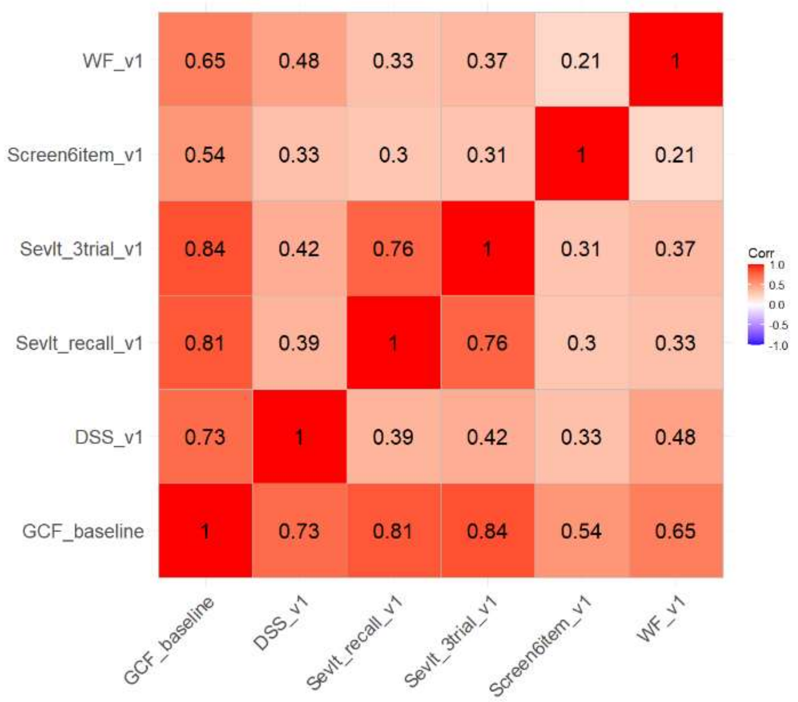


**Panel B. MESA Baseline (Visit 5)**

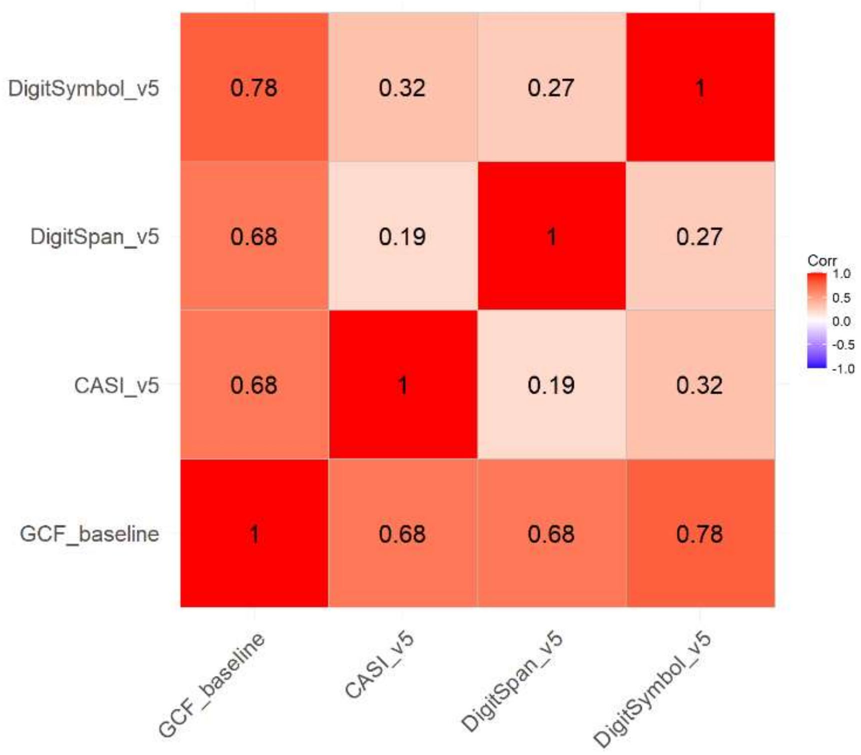

**Panel C. ARIC (Visit 4)**

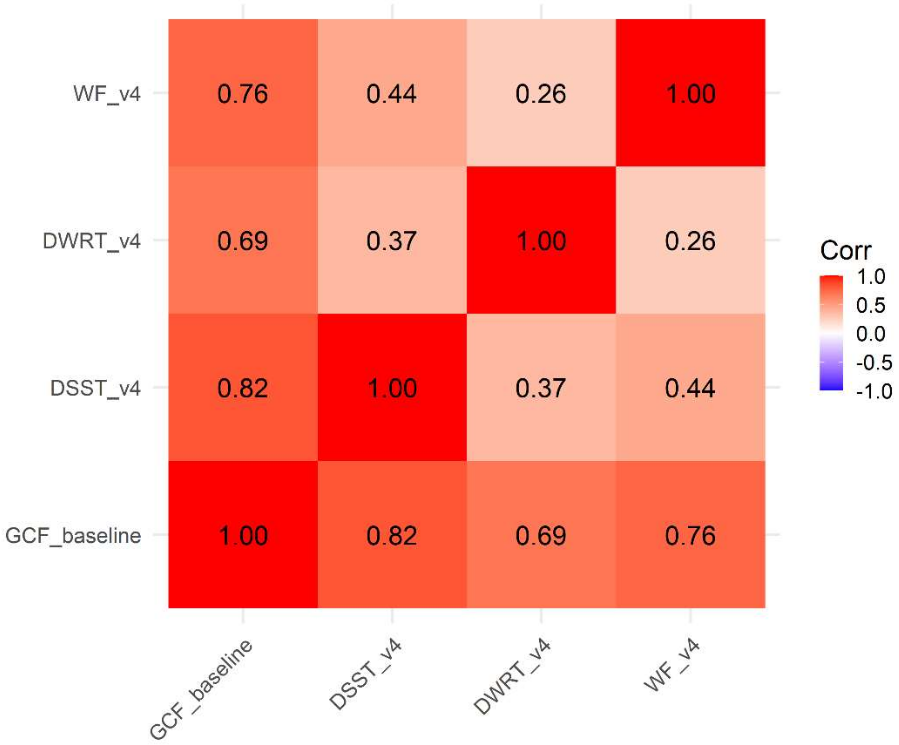

**Supplemental Figure 4. Study-Specific and Meta-Analysis Results for the Association between Continuous Sleep Phenotypes and Global Cognitive Function Measures**

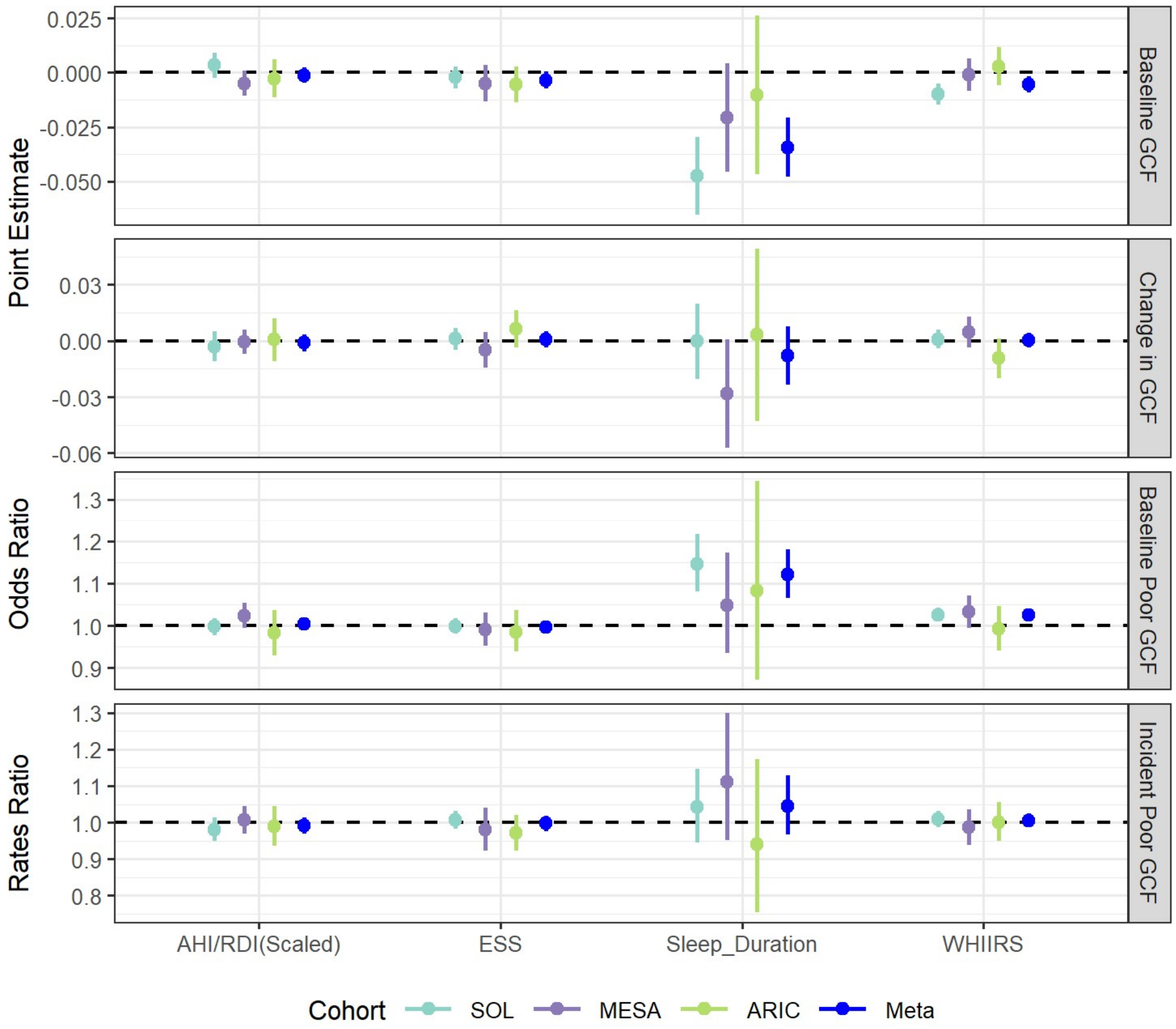

**Supplemental Figure 5. Sex-Stratified Study-Specific and Meta-Analysis Results for the Association between Sleep Phenotypes and Global Cognitive Function Measures**

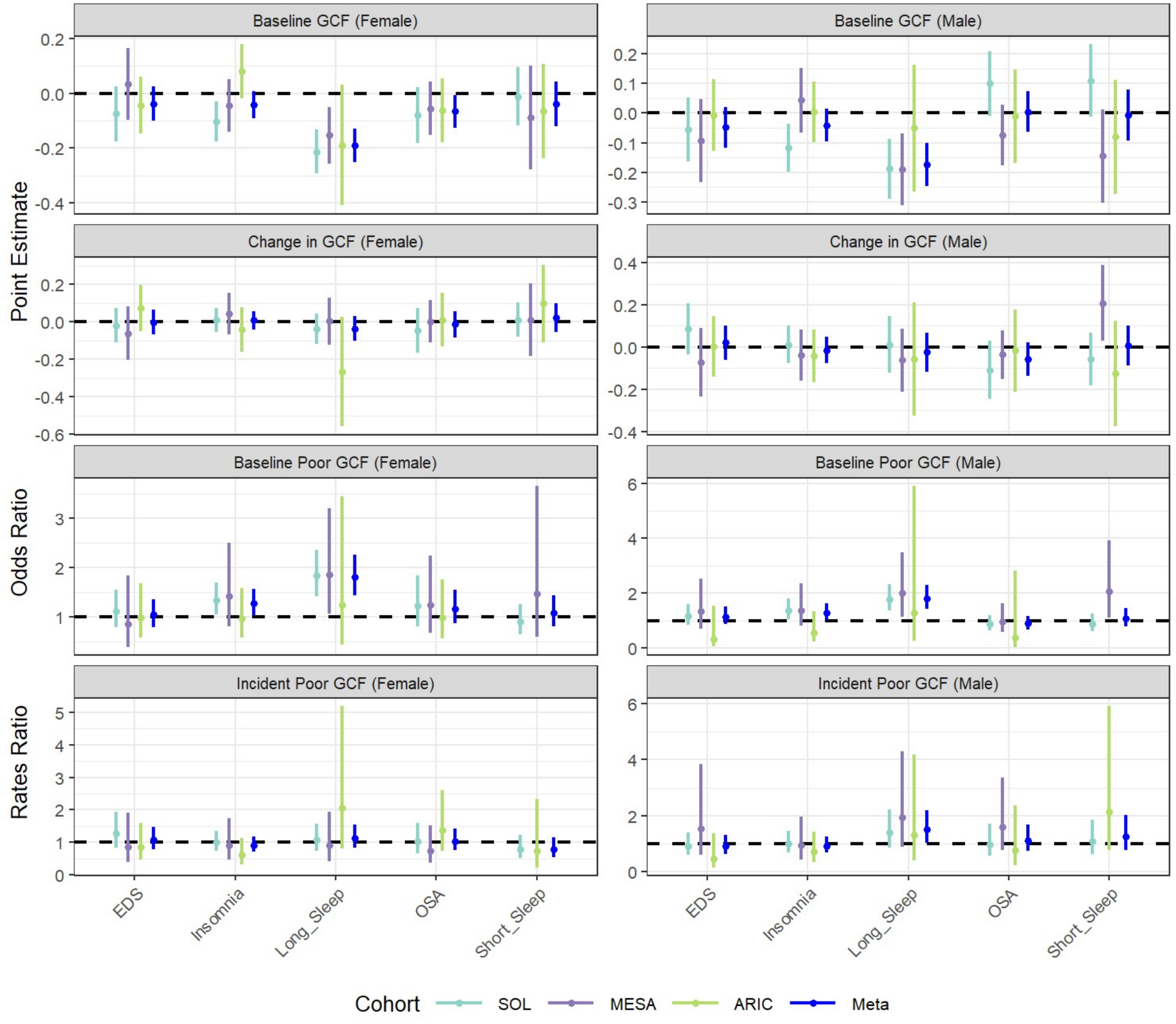

**Supplemental Figure 6. Study-Specific and Meta-Analysis Results for the Association between Sleep Phenotypes and Global Cognitive Function Measures using Inverse Probability Weighting (IPW)**

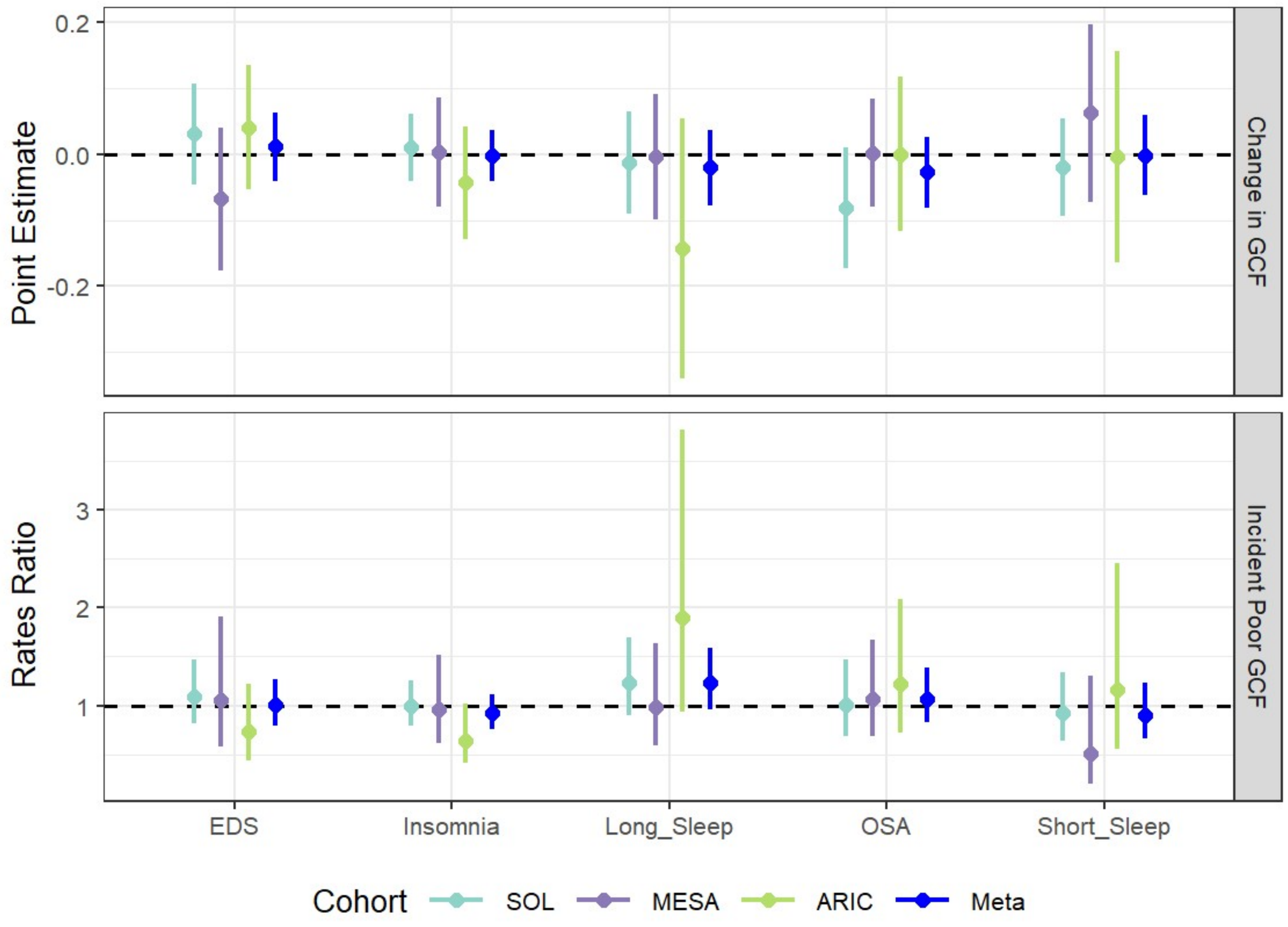
